# First trimester antidepressant use and miscarriage: a comprehensive analysis in the UK Clinical Practice Research Datalink

**DOI:** 10.1101/2024.10.19.24315779

**Authors:** Florence Z. Martin, Paul Madley-Dowd, Viktor H. Ahlqvist, Gemma C. Sharp, Kayleigh E. Easey, Brian K. Lee, Abi Merriel, Dheeraj Rai, Harriet Forbes

## Abstract

**Objectives:** To investigate the risk of miscarriage associated with first trimester antidepressant use.

**Design:** Population-based cohort study.

**Setting:** UK Clinical Practice Research Datalink (CPRD) GOLD.

**Participants:** 661 825 individuals who had 1 021 384 pregnancies in CPRD GOLD between 1996 and 2018.

**Main outcome measures:** Miscarriage defined as a pregnancy loss prior to 24 weeks’ gestation.

**Results:** Among the eligible pregnancies, 73 540 were prescribed antidepressants in trimester one (7.2%); 14.7% antidepressant prescribed pregnancies ended in miscarriage, as opposed to 12.4% of those not prescribed antidepressants. Antidepressant use during trimester one was associated with miscarriage in the unadjusted models (hazard ratio (HR) 1.21, 95% confidence interval (CI) 1.19 to 1.23), which attenuated following adjustment for covariates (aHR 1.04, 95% CI 1.02 to 1.06). These findings translated to an absolute risk adjusted for confounders of 13.1% (95% CI 13.0 to 13.2) in the unexposed compared to 13.6% (95% CI 13.3 to 13.8) in the first trimester antidepressant exposed. The propensity score matched model showed similar results (aHR 1.09, 95% CI 1.02 to 1.17, respectively). In those with depression or anxiety in the 12 months before pregnancy, our estimate didn’t change (aHR 1.04, 95% CI 1.01 to 1.08).

**Conclusion:** First trimester antidepressant use was associated with a small yet clinically insignificant increase in risk of miscarriage, with no evidence suggesting taking antidepressants before pregnancy and into first trimester increases the risk of miscarriage. The conclusions are less clear for ‘incident’ antidepressant use in trimester one, however issues including gestational dating in early pregnancy and probable residual confounding prohibit us from interpreting this observation as causal.

## 1 INTRODUCTION

Antidepressant use during pregnancy is prevalent in many countries, with estimates suggesting that upwards of 8% of pregnant people in the United Kingdom,^1^ Iceland,^2^ and the United States^3^ use antidepressants at some point during pregnancy. Although most antidepressants are not contraindicated during pregnancy, they are prescribed with some caution,^4^ due to evidence suggesting small increases in risk of miscarriage^5–7^ and other adverse outcomes, such as preterm delivery and postpartum haemorrhage.^8^ ^9^ In the UK, the National Institute for Health and Care Excellence (NICE) updated its guidance in 2023 from severity-based advice to patient-centred decision-making when planning pregnancy or becoming pregnant on antidepressants, weighing up risks to both mum and baby on an individual basis.^10–12^ Globally, the guidance around using antidepressants during pregnancy is mixed,^13^ reflecting the uncertainty in the evidence base and in turn, challenges faced by prescribing clinicians.

The definition of miscarriage varies in different countries and time periods, but is often defined as a pregnancy loss before 20–24 weeks’ gestation.^14^ ^15^ A systematic review and meta-analysis of 29 studies identified a modest increased risk of miscarriage following any antidepressant use during pregnancy (pooled odds ratio 1.24, 95% confidence interval (CI) 1.18 to 1.31).^16^ Biologically, it is plausible that antidepressants could causally increase the risk of miscarriage, due to their inhibition of platelets and subsequent association with increased bleeding events.^17^ However, untreated depression and anxiety during pregnancy are also associated with adverse pregnancy outcomes, like preterm birth and low birthweight.^18–21^ Thus, it is plausible that the link between antidepressant use during pregnancy and adverse outcomes like miscarriage, could be explained by the underlying disease for which antidepressants are prescribed,^1^ ^22^ rather than the drugs themselves; this concept is known as confounding by indication. Given the use of general population, non-indicated controls in many of the included studies in the above systematic review,^16^ and some studies omitting adjustment for underlying reason for prescribing,^23–29^ it isn’t possible to conclude a causal relationship between antidepressants and miscarriage from the present literature.

In this cohort study, we used Clinical Practice Research Datalink (CPRD) GOLD data, with linked Hospital Episode Statistics (HES) data where available, to investigate the relationship between first trimester use of antidepressants and the risk of miscarriage using a range of advanced methodological approaches, including an exposure discordant pregnancy design, propensity score matching, and stratified analyses, to help account for confounding by and severity of the underlying disease.

## 2 METHODS

### 2.1 DATA SOURCES

CPRD GOLD is a UK-wide repository of anonymised general practice data and makes up part of one of the largest resources of primary care data in the world.^30^ It contains over 4 million active patients and covers ∼7% of the UK’s population, representative by age, sex, and ethnicity.^30^ The primary care data in CPRD GOLD is linked to the Office for National Statistics death registration data and practice-level Index of Multiple Deprivation (IMD) scores. For most English patients, CPRD GOLD is linked with Hospital Episode Statistics (HES) Admitted Patient Care (APC), covering inpatient hospital episodes.^30^

The CPRD GOLD Pregnancy Register has been described in detail elsewhere,^31^ but in short, it is a dataset that contains pregnancy episodes with affiliate estimated pregnancy dates, outcome, and patient identifiers derived from the CPRD GOLD primary care data. The data sources used in this study are detailed in Methods S2.1.

### 2.2 STUDY POPULATION

To derive the study population, eligibility criteria were imposed on the entire CPRD GOLD population who had a record of at least one pregnancy episode between 1996 and 2018 in the Pregnancy Register. We cleaned the Pregnancy Register in accordance with recommendations from the authors of the Register algorithm, including removing conflicting and historical pregnancies.^32^

HES data were used to supplement pregnancy outcomes that were uncertain in the Pregnancy Register (namely ‘unknown outcome’ and ‘unspecified loss’). Pregnancy dates in the Pregnancy Register were then amended using imputed values as imposed by the Pregnancy Register algorithm (Methods S2.2). Pregnancy episodes ending in an ‘unknown outcome’ that were not recoverable using HES were excluded.

CPRD imposes an ‘up-to-standard’ (UTS) date on all enrolled practices, which records the date on which the practice began to contribute ‘high quality’ data to CPRD, defined by several indicators.^30^ We only included those who were registered with a UTS practice and had adequate follow-up for at least a year prior to pregnancy and up until the end of pregnancy; by extension, eligible individuals did not transfer out of their practice or have a death date recorded prior to the end of pregnancy. Individuals were also excluded if the last data collection date from their practice occurred before the end of their pregnancy.

### 2.3 EXPOSURE

All antidepressants that are approved for treating depression in the UK were extracted from primary care prescriptions (Table S1). Briefly, quantity (total number of tablets prescribed) and daily dose (number of tablets taken per day) were used in conjunction with the prescription start date to estimate the prescription end date (Methods S2.3). These dates were compared with the pregnancy start date and the end date of trimester one to identify whether a prescription occurred within or overlapped with the first trimester to identify ‘exposed’.

Those with a prescription for antidepressants in the three months before pregnancy and during trimester one were defined as ‘prevalent’ users. Those without antidepressants in the three months prior to pregnancy but prescribed during trimester one were defined as ‘incident’ users.

We identified antidepressant class prescription in trimester one: selective serotonin reuptake inhibitors (SSRI), tricyclic antidepressants (TCA), serotonin-noradrenaline reuptake inhibitors (SNRI), ‘other’ (Table S1), or multiple classes (i.e., those who switched product class during trimester one or used multiple antidepressants from different classes simultaneously).

Finally, we defined dose, standardised for each medication using the individual dose distribution in milligrams and using percentiles to ascertain low (≤25^th^ percentile), medium, and high (>75th percentile) doses (Methods S2.3). In instances where multiple doses were prescribed in trimester one, individuals were classified as the highest dose they received in first trimester.

### 2.4 OUTCOMES

The outcome of each pregnancy episode was available in the Pregnancy Register.^31^ Miscarriage from the Pregnancy Register was used as the outcome in this study.

### 2.5 COVARIATES

Confounders were chosen based on subject matter knowledge of whether a covariate could feasibly cause both the exposure and the outcome. The primary adjustment set contained age, year of pregnancy (‘96–’00, ‘01–’05, ‘06–’10, ‘11–’15, ‘16–’18), IMD quintile, history of miscarriage and severe mental illness, smoking (non-, ex-, current smoker), parity (0, 1, 2, ≥3), use of high dose folic acid, antipsychotics and anti-seizure medications in the 12 months before pregnancy, number of primary care consultations in the 12 months before pregnancy (0, 1–3, 4– 10, >10), and depression and anxiety ever before the start of pregnancy; this is described detail in Methods S2.5.

Depression and anxiety were identified using pre-defined, expert verified codelists in primary care (Read codes) and HES APC (ICD-10 codes) (Methods S2.5).

Ethnicity (White, South Asian, Black, Other, Mixed)^33^ and body mass index (BMI; <18, 18–24.9, 25–29.9, >30kg/m^2^) around the start of pregnancy contained >10% missing data; thus, they were dropped from the primary adjustment set and included in a sensitivity analysis, described below.

### 2.6 ANALYSIS

We described baseline characteristics of the eligible sample by first trimester antidepressant use. We also compared the characteristics between eligible pregnancies and those excluded having ended in ‘unknown outcome’. We ran the primary and secondary analyses in complete records for covariates.

#### 2.6.1 PRIMARY ANALYSES

##### 2.6.1.1 MULTIVARIABLE COX MODEL

We compared antidepressant exposed to unexposed, using crude and adjusted Cox models estimating hazard ratios (HR) and 95% CIs. Follow-up began on the estimated pregnancy start date; ‘incident’ users contributed unexposed time to the analysis until the start of their antidepressant prescription. The end of follow-up was set to the first of either the outcome (miscarriage), other loss (Table S4), reaching 168 days gestation, or study end (31^st^ December 2018). We employed cluster-robust standard errors (clustered by pregnant individual) to account for those who contributed multiple pregnancies to the analysis.

To enhance clinical interpretability, we estimated the absolute confounder-adjusted risks (1-Survival) using Breslow’s baseline estimator and integrated these with the hazard ratios through G-formula and bootstrapping for standard errors (1000 repetitions).

In addition to adjusting for indication, we ran the model restricted to those with evidence of depression or anxiety in the 12 months prior to pregnancy. To further investigate severity, we restricted the model to those with ‘severe’ depression or anxiety, as defined by administered scale standardised scores (like PHQ-9, Methods S2.6) in the 12 months before pregnancy.

We additionally restricted the sample to those who were prescribed antidepressants in the three months prior to pregnancy; we compared those who continued antidepressants into trimester one to those who discontinued treatment prior to the start of pregnancy. This assumed that those using antidepressants pre-pregnancy were more characteristically similar, thus lessening residual confounding.

##### 2.6.1.2 EXPOSURE DISCORDANT PREGNANCIES

We held genetic liability to miscarriage and variables that were not time-varying between pregnancies fixed by comparing pregnancies among the same individual in an exposure discordant design as an additional approach to manage confounding.^34^ ^35^ We used a stratified Cox model adjusted for the primary adjustment set (except history of miscarriage), where each stratum in the model represented an individual with ≥2 exposure discordant pregnancies (Methods S2.6).

##### 2.6.1.3 PROPENSITY SCORE MATCHING

We performed propensity score matching, following the stepwise process laid out by Desai *et al*.^36^ The propensity score included both putative confounders and predictors of the outcome (Table S2).^37^

The sample in the propensity score matched analysis only included first pregnancies, to avoid individuals being matched to their own subsequent pregnancies. We used logistic regression to estimate a propensity score, then 1:1 matched each antidepressant exposed pregnancy to an unexposed pregnancy without replacement using a caliper of 0.2, and exact matching on number of CPRD consultations in the 12 months before pregnancy (Methods S2.6).

#### 2.6.2 SECONDARY ANALYSES

##### 2.6.2.1 ‘PREVALENT’ AND ‘INCIDENT’ ANALYSIS

‘Prevalent’ and ‘incident’ antidepressant users were compared to unexposed. We also performed this analysis among those with any depression or anxiety and ‘severe’ illness (Table S3) in the 12 months before pregnancy.

##### 2.6.2.2 CLASS AND DOSE ANALYSES

We compared different antidepressant classes to unexposed. We also compared low, medium, and high doses of antidepressant in trimester one to no use (Methods S2.3).

#### 2.6.3 SENSITIVITY ANALYSES

We restricted the primary and secondary analyses to those with linked secondary care data, due to pregnancy outcome modifications and high data availability in this group (Methods S2.2). We also performed the primary Cox model where exposure was redefined as ≥2 antidepressant prescriptions in trimester one to reduce potential exposure misclassification.

We investigated the association between pre-pregnancy depression, anxiety, antidepressant use and having a pregnancy that ended in an ‘unknown outcome’ to assess the potential for differential pregnancy exclusion from the sample.

Having dropped ethnicity and BMI from the adjustment set due to >10% missing data, we included these covariates in sensitivity analysis. We also investigated the association between missingness in these variables and experiencing a miscarriage to assess the potential introduction of bias in the complete records analysis.^38^

To account for the potential effect of behavioural changes between pregnancies where the outcome of one pregnancy influences care seeking and provision in the next pregnancy, we restricted the primary Cox model to first pregnancies.

To test the impact of censoring pregnancies that ended in other types of early pregnancy loss, including ectopic and molar pregnancies (Table S4), we absorbed them into the miscarriage category in sensitivity analysis.

All analyses were performed in Stata 17 and R 4.3.1. This study was approved by CPRD’s Independent Scientific Advisory Committee (ISAC) in 2021 [ISAC number: 21_000362].

### 2.7 PATIENT AND PUBLIC INVOLVEMENT

No patients were directly consulted regarding the definition of the research question, study design, analyses, or write-up. We shared our plans at public engagement events, including the Pint of Science festival.^39^ We consulted with clinical colleagues and the British Pregnancy Advisory Service (BPAS) who are in regular discussion with pregnant people concerned about the risk of miscarriage following first trimester antidepressant use. This provided sufficient motivation for the importance of the present study to individuals of child-bearing age considering antidepressant treatment.

## 3 RESULTS

### 3.1 STUDY POPULATION

The CPRD GOLD Pregnancy Register contained 1 245 146 non-conflicting pregnancies that began between 1996 and 2018 with sufficient follow-up. Upon the exclusion of ‘unknown outcome’ and multiple pregnancies, 1 021 384 pregnancies (among 661 825 individuals) were eligible (Figure 1). Pregnancy outcomes in the eligible sample are summarized in Table S4.

**Figure 1.**
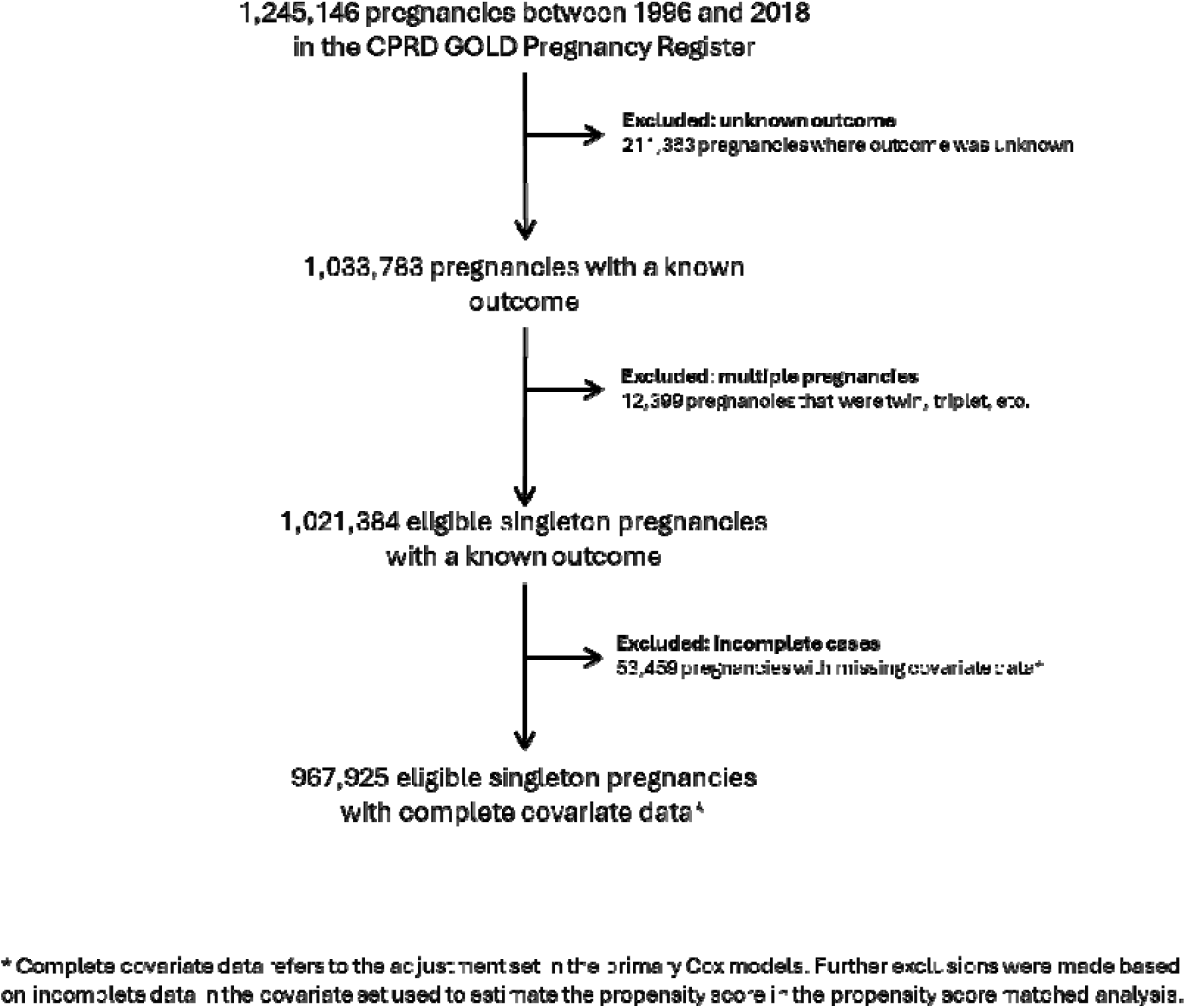
Sample selection and flow of pregnancy episodes through the study.

### 3.2 POPULATION CHARACTERISTICS

Following exclusions, 73 540 were prescribed antidepressants during trimester one (7.2%). Those prescribed antidepressants during trimester one were slightly older (22.7% *v* 19.7% over 35) and were more likely to be obese (25.2% *v* 17.0%) than unexposed. Individuals prescribed antidepressants during trimester one were more likely to be from the most deprived IMD quintile (30.7% *v* 26.9%) than unexposed. Those prescribed antidepressants were more likely to visit their doctor over 10 times (53.7% *v* 19.1%) and be using other medications (e.g., mood stabilisers, 4.1% *v* 0.7%) in the 12 months prior to pregnancy, be multiparous, and be current smokers than unexposed (Table 1).

**Table 1.**
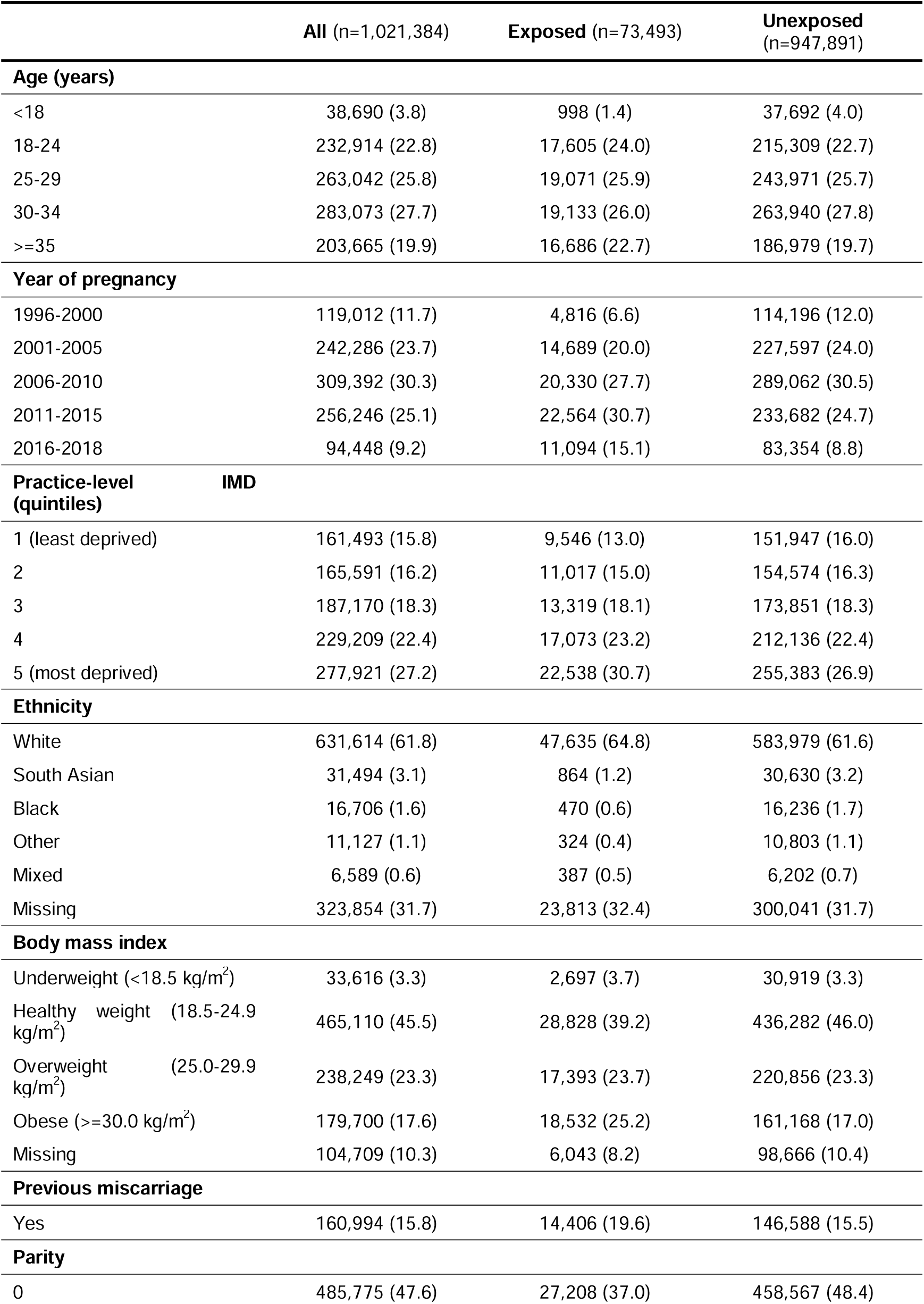
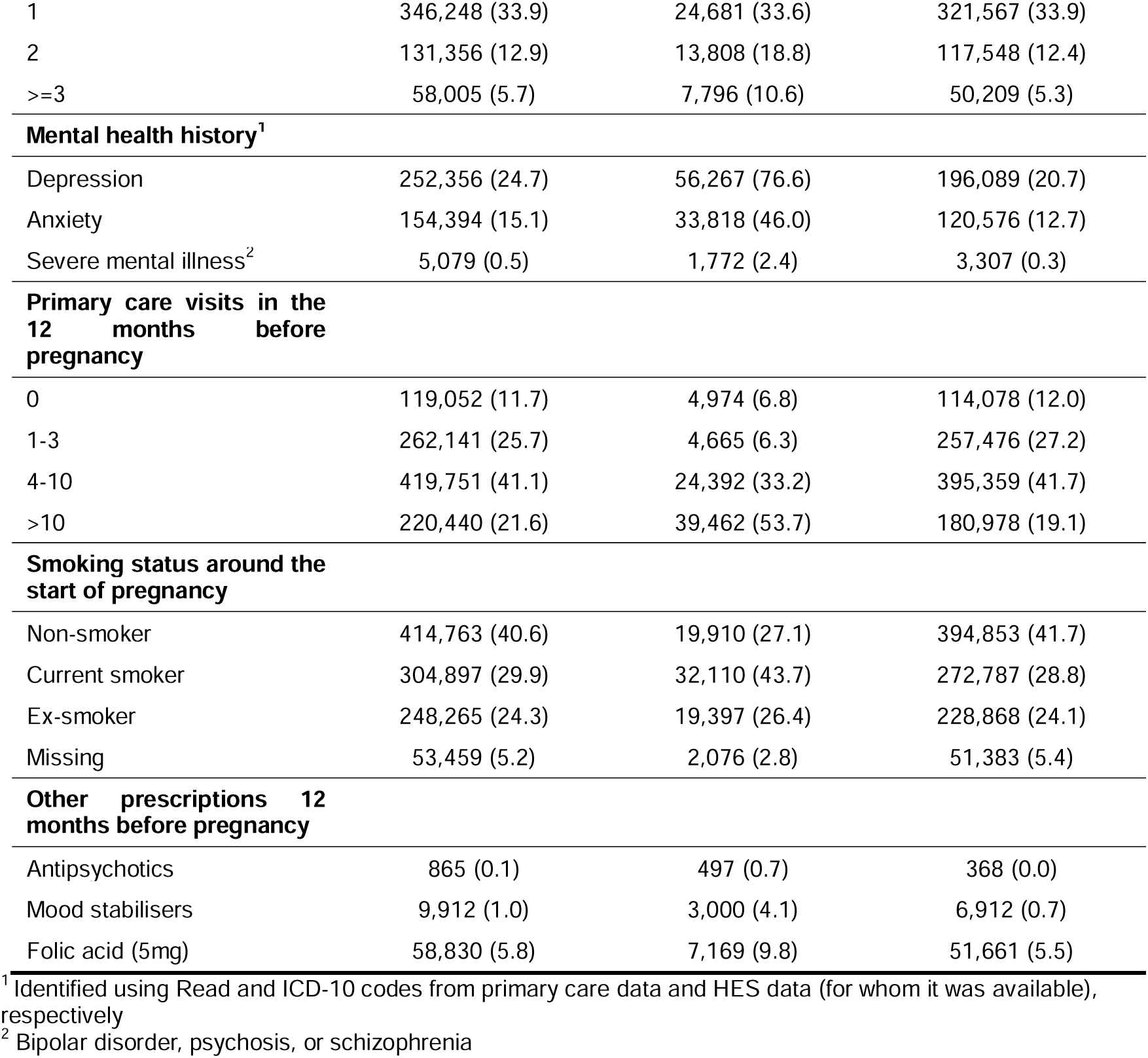
Characteristics of pregnant women eligible for inclusion.

Those excluded due to an ‘unknown outcome’ pregnancy were broadly similar to the eligible individuals, other than higher amounts of missing data in certain variables and on average more doctor visits before pregnancy (Table S5).

### 3.3 PRIMARY COX MODEL

Among 967 925 complete record pregnancies, 71 460 were exposed to antidepressants in trimester one, with 14.6% ending in miscarriage, compared to 12.3% of the 896 465 unexposed pregnancies (unadjusted HR 1.21, 95% CI 1.19 to 1.23). Upon adjustment, the difference between groups decreased (adjusted HR (aHR) 1.04, 95% CI 1.02 to 1.06), with a standardized miscarriage risk of 13.6% (95% CI 13.3 to 13.8) in the exposed group and 13.1% (95% CI 13.0 to 13.2) in the unexposed group (Figure 2). This finding was consistent when we required ≥2 distinct antidepressant prescriptions in trimester one to be considered exposed (aHR 1.02, 95% CI 1.00 to 1.05) (Table S6).

**Figure 2.**
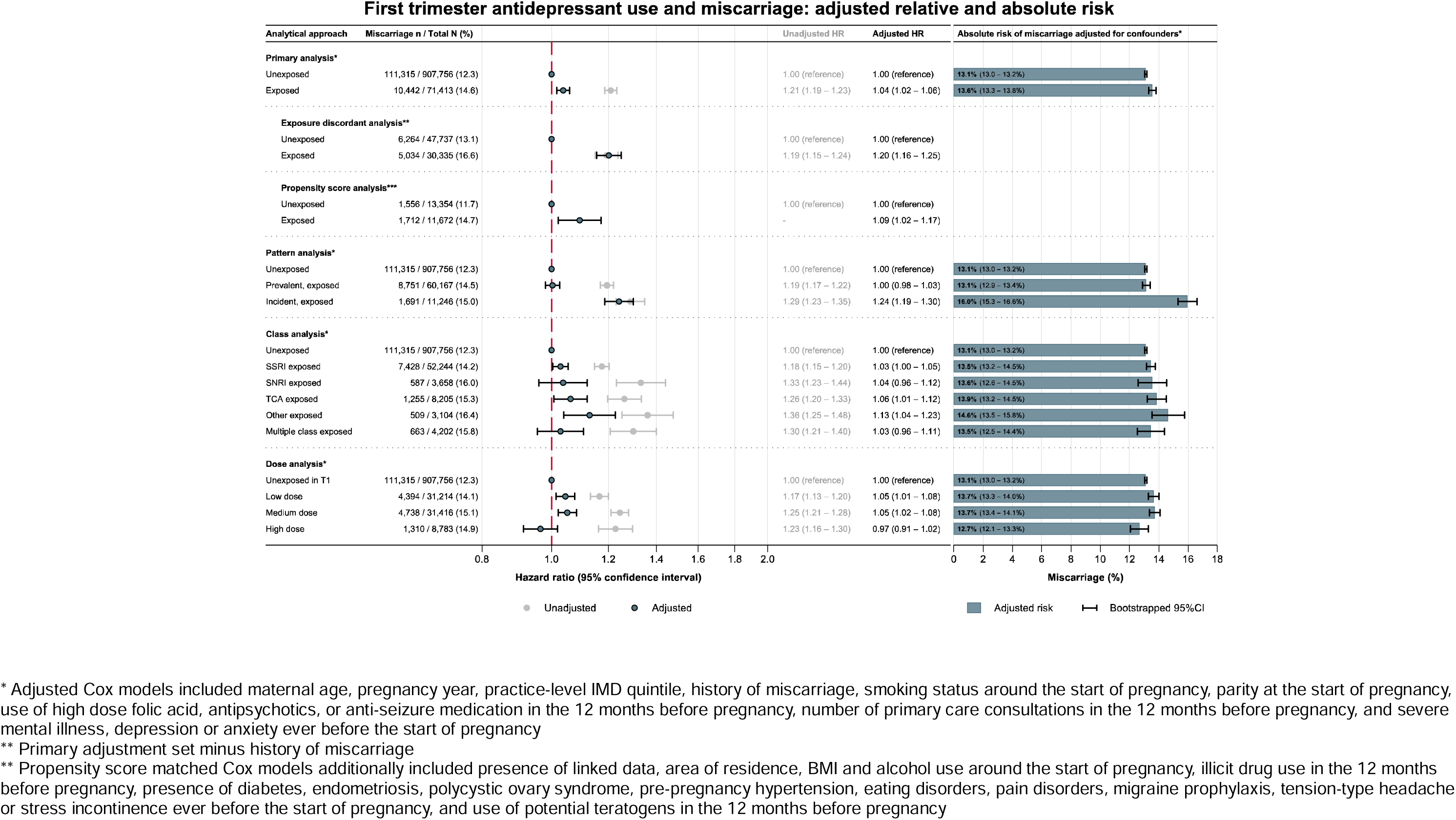
Findings from the primary and secondary analyses.

When restricting to those with depression or anxiety noted in the 12 months prior to pregnancy (n=99 820) and those with “severe” depression (n=9170), we observed similar results to the primary analysis (aHR 1.04, 95% CI 1.01 to 1.08 and aHR 1.02, 95% CI 0.92 to 1.14, respectively) (Table S7).

When comparing those who continued antidepressants into trimester one (n=60 167) to those who discontinued in the three months before pregnancy (n=24 410), we observed no difference in hazard of miscarriage (aHR 1.00, 95% CI 0.97 to 1.04) (Table S8).

### 3.4 EXPOSURE DISCORDANT PREGNANCY ANALYSIS

When comparing exposure discordant pregnancies within the same birthing parent (n=78 072), thereby accounting for all unobserved (e.g., genetics and many environmental factors that don’t change between pregnancies within an individual) and observed stable confounders, we saw an effect in line with the unadjusted primary Cox model (aHR 1.20, 95% CI 1.16 to 1.25) (Figure 2).

To understand whether this may have been driven by order of pregnancy, we investigated the risk of miscarriage when the first pregnancy in the exposure discordant group of pregnancies was exposed and then when a subsequent pregnancy in the group was exposed. This revealed that first trimester antidepressant use was only associated with miscarriage when the first pregnancy in the group was exposed (aHR 1.98, 95% CI 1.82 to 2.16), not when a subsequent pregnancy was exposed (aHR 0.97, 95% CI 0.93 to 1.02) (Figure 3, Table S10). The stark difference between these results suggests that pregnancy order may be driving the result observed in the exposure discordant analysis as opposed to the medication itself.

**Figure 3.**
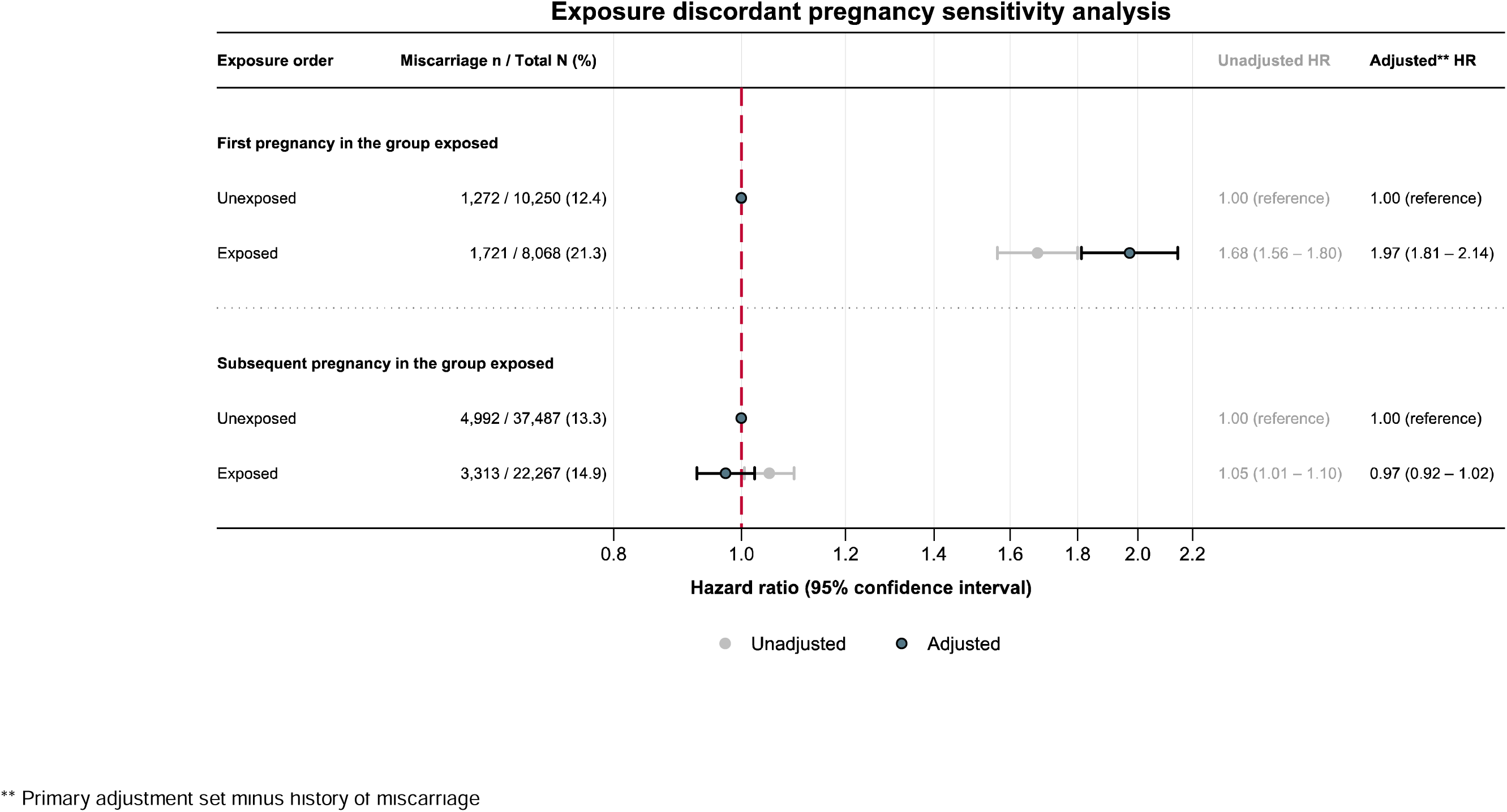
Exposure discordant pregnancy sensitivity analysis, restricting first to exposure discordant groups where the first pregnancy was antidepressant exposed and subsequent pregnancies in the group were not, then to groups where subsequent pregnancies were exposed to antidepressants but first pregnancies in the group were not.

### 3.5 PROPENSITY SCORE MATCHING

When matching pregnancies on propensity score (n=25 026) where more measured confounders were accounted for, our findings were consistent with those from the primary Cox model (aHR 1.09, 95% CI 1.02 to 1.17) (Figure 2).

### 3.6 ‘PREVALENT’ AND ‘INCIDENT’ ANALYSIS

In the unadjusted models, both ‘prevalent’ and ‘incident’ use in trimester one was associated with an increased hazard of miscarriage (HR 1.19, 95% CI 1.17 to 1.22 and HR 1.29, 95% CI 1.23 to 1.35, respectively). Interestingly, adjustment for covariates only changed our conclusions for ‘prevalent’ use, not ‘incident’ use (aHR 1.00, 95% CI 0.98 to 1.03 and aHR 1.24, 95% CI 1.19 to 1.30, respectively) (Figure 2), despite similarity across their measured characteristics (Table S9). Our conclusions did not change when restricting to those with ≥2 prescriptions in trimester one or when depression or anxiety were noted in the 12 months before pregnancy (Table S6, Table S7).

### 3.7 CLASS AND DOSE ANALYSIS

SSRI, SNRI, and ‘other’ antidepressant use during trimester one were associated with a slight increase in risk of miscarriage as compared to no use (Figure 2). Low and medium dose were associated with miscarriage, where high dose attenuated to the null as compared to unexposed following adjustment for covariates (Figure 2).

### 3.8 SENSITIVITY ANALYSES

Our results were consistent when restricting each analysis to those with linked data (Table S12). Having depression noted in the 12 months before pregnancy was modestly associated with having an ‘unknown outcome’ pregnancy (Table S13).

When adding ethnicity and BMI to the adjustment set for the primary Cox model, our estimates didn’t change (aHR 1.03, 95% CI 1.01 to 1.06) (Table S14). When assessing whether the complete records analysis may have been biased,^38^ those who had a miscarriage were more likely to have missing data in ethnicity, BMI, and smoking around the start of pregnancy (Table S15).

Similarly, when we restricted to first pregnancies, our estimates didn’t change substantially (aHR 1.07, 95% CI 1.03 to 1.10) (Table S16). When including ectopic and molar pregnancies into the definition of the outcome, the results were consistent with the primary analysis (aHR 1.03, 95% CI 1.01 to 1.05) (Table S17).

## 4 DISCUSSION

### 4.1 PRINCIPAL FINDINGS

This large population-based cohort study of nearly one million pregnancies in the UK found no clear evidence that first trimester antidepressant use substantially increases the risk of miscarriage, with no evidence suggesting taking antidepressants before pregnancy and into trimester one increases the risk of miscarriage. The conclusions are less clear for ‘incident’ first trimester antidepressant use, however issues including gestational dating in early pregnancy and probable residual confounding prohibit us from interpreting this observation as causal. The small observed increases in absolute risk, even if causal, are potentially clinically insignificant. The findings from the exposure discordant pregnancy analysis point to the importance of pregnancy order.

### 4.2 PREVIOUS LITERATURE

Previous literature exploring antidepressant use during pregnancy has suggested a slight increased risk of miscarriage, as shown by a systematic review and meta-analysis of 29 studies by Smith *et al*. which noted a number of methodological weaknesses in the previous literature.^16^ A large Danish study found an association between SSRI use during pregnancy and trimester one miscarriage of a similar magnitude to the findings presented here.^6^ They conclude that unmeasured lifestyle factors and confounding by indication are responsible for the small remaining association between SSRIs and miscarriage, given that they observed a complete attenuation of the effect to the null when comparing antidepressant exposure to unmedicated depression during pregnancy.^6^ Another study highlighted the challenges faced in the field of pregnancy pharmacoepidemiology, particularly when attempting to deal with confounding by indication.^40^ It is plausible that those who with ‘active’ depression (for example), have a higher baseline risk of miscarriage than those who do not have depression. If this is not properly handled in analyses of antidepressants and miscarriage, there is likely to be residual confounding by severity of indication.

Studies typically have accounted for indication by conducting additional analyses comparing those on antidepressant treatment during pregnancy with those who have unmedicated depression; some have found a complete attenuation to the null,^6^ ^40^ whereas others have found a persistent risk of miscarriage following antidepressant use.^5^ ^41–43^ Some have compared medication classes to account for confounding by indication, whereby both the “exposure” and “comparator” groups are likely to have an indication for antidepressants because they’re all exposed to antidepressants. Three studies have leveraged the comparison between SNRIs and other antidepressants,^44–46^ whereby a pooled increased risk of miscarriage was observed for those taking SNRIs.^16^ However, SNRI antidepressants are not first-line treatments in the UK, thus those prescribed them during pregnancy are likely to be more unwell. Only four studies in the review included variables pertaining to indication in a multivariable model.^40^ ^46^ ^47^ This highlights the persistent problem of confounding by severity of indication that is rarely eliminated even when accounting for indication; furthermore, those that use an indication-based sample may be prone to bias amplification.^48^ It is interesting that the unadjusted estimate from the present study, HR 1.21 (1.19-1.23), is similar to the summary estimate observed in the above review: pooled OR 1.24 (1.18 to 1.31).^16^

We show some novel findings surrounding ‘incident’ use of antidepressants during trimester one and miscarriage, where we observed a higher risk compared to no use and the confidence intervals do not overlap with the ‘prevalent’ use *v* no use. This intriguing association of ‘incident’ but not ‘prevalent’ use of antidepressants has been observed previously for some neurodevelopmental outcomes.^49^ Although these findings could be causal, whereby the introduction of a new drug substance into the body could disrupt early fetal development and result in an early pregnancy loss, there are several other plausible explanations that could explain the finding. As discussed above, residual confounding by severity of indication, health-seeking behaviour, or data artefacts like the imputation of pregnancy length for most losses might be partially driving the association. Antidepressant initiation symptoms such as heightened anxiety^50^ and the ongoing experience of symptoms during the time taken for antidepressants to start working^51^ may present alternative mechanisms for an increased risk of miscarriage that should also be considered when interpreting the finding for ‘incident’ users.

### 4.3 STRENGTHS AND WEAKNESSES

This study has several strengths. It is large, with over 600 000 individuals from a UK-representative sample,^30^ contributing nearly one million pregnancies over two decades, improving precision of our estimates. It leverages multiple methods and comparators to explore the role of confounding by indication and data issues encountered when performing observational pregnancy pharmacoepidemiology studies, particularly in early pregnancy loss. The use of the CPRD GOLD Pregnancy Register allowed us to build on the systematic approach taken by Minassian *et al.* that extracted pregnancy records from individuals who had been pregnant in the CPRD GOLD database.^31^ The use of cause-specific time-to-event models allowed us to retain pregnancies that were at risk of miscarriage while ongoing, but neither ended in the outcome nor reached the end of follow up, i.e., had ended in a non-miscarriage loss before week 24. It is important to consider the impact of their inclusion here; by keeping them in, we did not differentially deflate the denominator by exposure status and thus artificially inflate the proportion of pregnancies among the exposed group that ended in miscarriage. This omission from previous studies may have partially driven reports of an increased risk of miscarriage following use of antidepressants, even among studies that had adjusted for confounders.

The study also has several limitations. Although the CPRD GOLD population is large, the application of eligibility criteria based on registration in a UTS practice and quality of patient data inevitably led to a smaller and more select sample of individuals. We can be reassured that those excluded for having an ‘unknown outcome’ were similar characteristically to those included, but the findings likely only generalise to those that fulfil the criteria for this study, namely staying with the same practice for a year before and throughout pregnancy.

Residual confounding is likely present in these analyses despite our mixed approaches to accounting for it. Although we managed confounding by indication as completely as possible, like adjusting for depression and anxiety in the main analysis and restricting to those visiting the doctor for depression and/or anxiety or those having scored highly on depression and anxiety scales in the 12 months before pregnancy, it remains difficult to capture indication severity using CPRD. Thus, residual confounding by underlying severity of indication for treatment surely contributed to the results we observed, particularly for ‘incident’ use.

Systematic bias in these analyses cannot be ruled out. Differential exposure misclassification was a concern in this study as pregnancies ending in miscarriage were more likely to have an imputed gestational length than deliveries^31^ and therefore at higher risk of being misclassified as prescribed antidepressants in trimester one. The results may have been biased in either direction if this type of bias was present in these analyses.^52^ It is plausible that, given the imputation of gestational length for many losses, antidepressant prescriptions were sought having experienced a miscarriage. Due to the derivation of pregnancy dates via a pregnancy algorithm, the possibility for reverse causation may explain some of the miscarriages observed in the ‘incident’ group. Finally, ascertainment bias is likely at play here. Those seeking healthcare for depression, anxiety, or other indications treated with antidepressants may be more likely to report pregnancies and early pregnancy losses than those not engaging with healthcare for other reasons.

Given that the presence and magnitude of each of these limitations cannot be easily quantified, it is reassuring that even if the finding from the main analysis was causal, it would translate to a modest increase in absolute risk from 13.1% in the unexposed to 13.6% in the exposed (i.e., a number need to harm of 200).

### 4.4 FUTURE WORK

Where high quality miscarriage data are available, other causal inference approaches that aim to manage time-related biases like target trial emulation would be useful to explore the finding for ‘incident’ use. Given the issue of time in these data, where most miscarriages have an imputed gestational length, CPRD GOLD may not be appropriate for this. It is important to understand this finding to adequately inform individuals who may need to initiate antidepressants in the first trimester.

### 4.5 CONCLUSIONS

We found no clear evidence that antidepressant use during trimester one substantially increases the risk of miscarriage, with no evidence suggesting taking antidepressants before pregnancy and into trimester one increases the risk of miscarriage. Although we observed a slight increased risk of miscarriage when comparing ‘incident’ antidepressant use in trimester one to no use, the overall relative risk translates to a modest increase in absolute risk and other biases cannot be ruled out. Our findings suggest that antidepressants do not substantially increase the risk of miscarriage for women on antidepressants when they become pregnant.

## What is already known on this topic

- Antidepressant use during pregnancy was shown to increase the risk of miscarriage according to a recent systematic review.
- Confounding, including by indication, remains a pervasive problem in the interpretation of the current evidence.

## What this study adds

- A comprehensive analysis of first trimester antidepressant use and risk of miscarriage in the CPRD GOLD Pregnancy Register, including multiple approaches to address confounding.
- Estimates of standardised absolute risk of miscarriage among antidepressant exposed and unexposed to antidepressants in trimester one to aid in clinical interpretability of the findings.
- The results, particularly for ‘prevalent’ antidepressant use, are reassuring and support minimal risk of miscarriage following ongoing use of antidepressants into trimester one from pre-pregnancy.

## 5 ETHICAL STATEMENT

This study was approved by CPRD’s Independent Scientific Advisory Committee (ISAC) in 2021 [ISAC number: 21_000362].

## 6 DATA AVAILABILITY STATEMENT

“Access to CPRD data, including UK Primary Care Data, and linked data such as Hospital Episode Statistics, is subject to protocol approval” as per CPRD’s guidelines. Authors are unable to share the data in its raw form, however all analytical code and codelists are open-source and found via the following links: https://github.com/flozoemartin/Miscarriage and https://github.com/flozoemartin/codelists.

