## Supplementary material for "First trimester antidepressant use and miscarriage: a comprehensive analysis in the UK Clinical Practice Research Datalink": Methods S, Table S, Figure S

### 1 Contents

### 2 Methods

#### 2.1 Data source

The Clinical Practice Research Datalink (CPRD) is a primary care data repository in the UK that pulls data from the Vision (GOLD) and EMIS (Aurum) software used in general practice. For the present study, CPRD GOLD was used due to the availability of the Pregnancy Register at the time of data acquisition.

CPRD GOLD is made up of several datasets covering all elements of primary care delivery including prescriptions, diagnoses, referrals, and immunisations. Patients are linked to their records throughout these data using a pseudonymised patient identifier and also to external datasets like Hospital Episode Statistic (HES) and Index of Multiple Deprivation (IMD) data where available.

Linkage with practice-level IMD data and Office for National Statistics (ONS) death registration data is available for everyone in CPRD GOLD. Linkage with HES, including in- and out-patient hospital data, are only available for a proportion of the English practices (~75%).^1^

#### 2.2 Study population

­The Pregnancy Register as provided by CPRD contains information on each pregnancy episode for each patient who has experienced a pregnancy in primary care.^2^ The dataset contains information such as estimated pregnancy start and end dates, the estimated length of the pregnancy, the outcome of the pregnancy, and if there was evidence of preterm delivery, for example. A substantial proportion of the pregnancy episodes in the Pregnancy Register have an unknown outcome; these are explained in detail elsewhere, but in short, pregnancies without a known outcome were pregnancy episodes identified by the Pregnancy Register algorithm that did not have sufficient codes affiliated with them in order to be able to ascertain what the outcome might have been. Given that these made up a significant proportion of the pregnancies in the Pregnancy Register, they were cleaned before exclusion. The cleaning process of the pregnancies without a known outcome involved using the secondary care linked data to find pregnancy episodes and align them within patients to unknown outcome pregnancy records in the Pregnancy Register. Using outcome data in HES, pertaining to either a loss in hospital or a delivery, we were able to salvage some ‘unknown outcome’ pregnancies from exclusion by assigning them a new known outcome from the secondary data. The algorithms were developed by PMD and HF, amended by FZM, using the codelists developed and approach laid out by Campbell *et al.*^3^

#### 2.3 Exposure

The prescription data in CPRD is held within the Therapy file and consists of all prescriptions made to each patient in CPRD GOLD. In order to ascertain whether a prescription overlapped with pregnancy, we needed prescription start and end dates. Only prescription start date is available in the CPRD prescription data, thus we needed to derive the prescription end date using daily dose (the number of doses prescribed per day) and quantity (the number of doses given per prescription). These variables are provided by CPRD but require some post-hoc cleaning for unusual and missing values. Daily dose was supplemented with dose number multiplied by dose frequency and quantity was supplemented with pack size, where available. Where some or all of these values were missing, we used hot-decking imputation to fill in the gaps in the prescription data.^4^ The approach follows a modal approach with decreasing levels of specificity until all the gaps are filled: first, missing daily dose and quantity are filled in using the modal value of each variable from prescriptions of the same drug within the same patient. If no such data are available, then the modal value from the same prescriptions among all patients are taken, and so on until the least specific criteria, among the same class, is used to fill in the final missing values.

All antidepressants that fall under the N06A World Health Organisation Anatomical Therapeutic Chemical classification^5^ were investigated. Bupropion and antidepressants delivered in direct conjunction with antipsychotics were removed upon guidance from clinical co-authors as not prescribed for depression in the UK. They were loosely categorized into class, summarized in Table S1.

Table S1 Individual drug substances in each designated class of antidepressants.

| **Selective serotonin reuptake inhibitors (SSRIs)** | **Tricyclic antidepressants (TCAs)** | **(Serotonin-) noradrenaline reuptake inhibitors (SNRI/NRIs)** | **Other antidepressants** |
| --- | --- | --- | --- |
| Citalopram | Amitriptyline | Duloxetine | Agomelatine |
| Escitalopram | Amoxapine | Reboxetine | Isocarboxazid |
| Fluoxetine | Butriptyline | Venlafaxine | Mianserin |
| Fluvoxamine | Clomipramine |  | Mirtazapine |
| Paroxetine | Desipramine |  | Moclobemide |
| Sertraline | Dosulepin |  | Nefazodone |
|  | Doxepin |  | Phenelzine |
|  | Imipramine |  | Tranylcypromine |
|  | Lofepramine |  | Trazodone |
|  | Maprotiline |  | Tryptophan |
|  | Nortriptyline |  | Vortioxetine |
|  | Protriptyline |  |  |
|  | Trimipramine |  |  |

##### 2.3.1 Deriving dose

Dose was derived for each drug, by taking the dose prescribed per day and presented as daily dose in milligrams. Given that different medications are dosed differently, but all follow thresholds of low, medium, and high dosing, we wanted to standardize doses across each medication for comparability in the secondary analysis of dose.

To standardize dose, we plotted to distribution of dose for each medication and identified the dose value that represented each quartile of the distribution. We assigned low dose as the bottom 25% of the distribution, medium as the middle 50% of the distribution, and high as the top 75%. For drugs that are rarely prescribed or have few dose values that are prescribed in clinical practice, these thresholds had to be amended to more stringent upper and lower bounds, for example agomelatine required an upper threshold of 90% to capture true high dose in the high dose category. The spreadsheet showing the dose thresholds for each medication is attached in the Appendix. The doses assigned to each medication were checked for clinical agreement in EMC and with clinical coauthors.

#### 2.4  Outcomes

Pregnancy outcome is provided in the Pregnancy Register;^2^ for the present study we used pregnancies defined as ending in miscarriage for the outcome of interest, as per the Pregnancy Register and ‘unknown outcome’ pregnancies that were reclassified as miscarriage based on the above cleaning process (Methods S2.2).

#### 2.5 Covariates

Covariates were identified *a priori*, derived in the data, and included in the adjustment set for the primary Cox model and the exposure discordant pregnancy analysis if they had <10% missing data. The potential adjustment set contained pregnancy start year, maternal age at the start of pregnancy,^6-9^ maternal body mass index (prior to and as close to the start of pregnancy as possible),^8 9^ socioeconomic position (using practice-level IMD score in quintiles),^7 10^ parity,^7^ previous losses (miscarriage),^6 8 9^ number of face-to-face primary care consultations in the 12 months prior to pregnancy start,^11^ prescriptions of high-dose folic acid (5mg) in the 12 months before pregnancy start,^7^ antipsychotics^12^ or anti-seizure medications^13^ in the 12 months prior to pregnancy start, ethnicity,^14-16^ smoking,^6^ and history of severe mental illness,^17^ depression, and anxiety^18^ ever before the start of pregnancy (Figure S1).

Similarly, the covariates for the propensity score matched analysis were identified *a priori* and included the above covariates. Those included in the propensity score analysis were complete cases for all covariates. In addition to the covariates included in the adjustment set for the primary and exposure discordant Cox models, the model to estimate the propensity score included whether the patient had linked data, gynaecological problems (endometriosis^19^ and polycystic ovary syndrome),^20^ pre-pregnancy hypertension,^21^ diabetes and hypothyroidism,^6^ and exposure to alcohol,^6 8^ illicit drugs, and known or suspected teratogenic drugs in the 12 months before pregnancy (Figure S1 and Table S2).

Figure S1 Directed acyclic graph of the adjustment graph in the analysis of antidepressant use during trimester one and miscarriage.

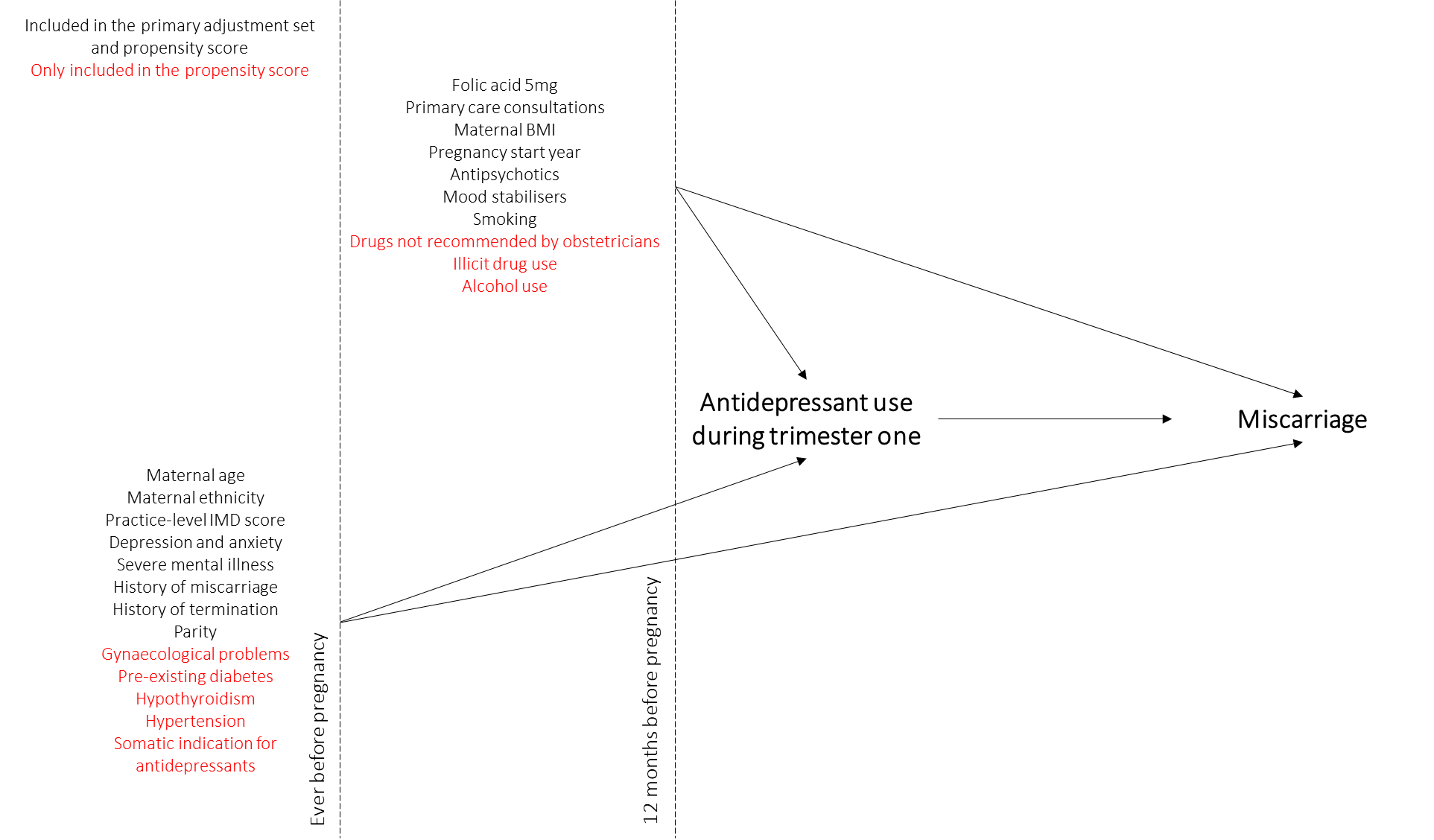

Arrows that exist between confounders to represent their relationships with each other have not been included here.

Table S2 Derivation and definition of each included covariate in the study.

| **Covariate** | **Definition^1^** |
| --- | --- |
| **Primary model** | |
| Maternal age | Age of mother in years at the start of pregnancy estimated from the Patient file in CPRD GOLD |
| Maternal ethnicity | Maternal ethnicity estimated using algorithmic approach developed by Mathur *et al.* ^22^ using the Clinical data in CPRD GOLD and HES Patient data, where available |
| Maternal body mass index (BMI) | Maternal BMI around the start of pregnancy derived hierarchically from the Clinical and Additional files in CPRD GOLD, prioritizing measures taken in trimester one |
| Pregnancy year | The year of pregnancy identified from the pregnancy start date in the CPRD GOLD Pregnancy Register |
| Parity | Number of previous deliveries within each woman in the Pregnancy Register (live birth, stillbirth, live birth or stillbirth, delivery based on a late record, delivery based on a third trimester record) |
| History of miscarriage | Whether any of the previous pregnancy episodes in each woman in the Pregnancy Register ended in miscarriage |
| Smoking status | Maternal smoking status around the start of pregnancy derived hierarchically from the Clinical and Additional files in CPRD GOLD, prioritizing measures taken during pregnancy |
| Practice-level Index of Multiple Deprivation (IMD) | The quintile of IMD assigned to each practice by CPRD GOLD, available for all CPRD GOLD practices |
| Primary care consultations | Number of primary care consultations that involved interaction with a healthcare professional in the 12 months before pregnancy using the Consultation file |
| High dose folic acid use | Prescriptions for products containing at least 5mg of folic acid in the 12 months before pregnancy using the Therapy file |
| Antipsychotic use | Prescriptions for antipsychotics in the 12 months before pregnancy using the Therapy file |
| Anti-seizure medication use | Prescriptions for anti-seizure medications (a.k.a., mood stabilisers or antiepileptics) in the 12 months before pregnancy using the Therapy file |
| Depression | Evidence of depression ever before the start of pregnancy using Read codes against the Clinical and Referral files and ICD-10 codes against the HES Diagnosis data, where available |
| Anxiety | Evidence of anxiety ever before the start of pregnancy using Read codes against the Clinical and Referral files and ICD-10 codes against the HES Diagnosis data, where available |
| Severe mental illness | Evidence of bipolar disorder, schizophrenia, or psychosis ever before the start of pregnancy using Read codes against the Clinical and Referral files and ICD-10 codes against the HES Diagnosis data, where available |
| **Propensity score matched analysis** | |
| Linked data | Those with (75% of English patients) and without (the reminder of the English patients and all patients from the devolved nations) linked HES data |
| Area of residence | CPRD region recorded in the Practice file |
| Alcohol use around the start of pregnancy | Maternal alcohol use status around the start of pregnancy derived hierarchically from the Clinical and Additional files in CPRD GOLD, prioritizing measures taken as early one year prior to and up to one month into pregnancy |
| Illicit drug use | Evidence of illicit drug use in the 12 months prior to pregnancy using Read codes in the Clinical and Referral files and prescriptions for medications such as methadone in the Therapy file |
| Diabetes | Evidence of diabetes (excluding gestational) ever before the start of pregnancy using Read codes in the Clinical and Referral files |
| Hypertension | Evidence of hypertension (excluding gestational) ever before the start of pregnancy using Read codes in the Clinical and Referral files |
| Endometriosis | Evidence of endometriosis ever before the start of pregnancy using Read codes in the Clinical and Referral files and ICD-10 codes in HES Diagnosis data, where available |
| Polycystic ovary syndrome (PCOS) | Evidence of PCOS ever before the start of pregnancy using Read codes in the Clinical and Referral files and ICD-10 codes in HES Diagnosis data, where available |
| Potential teratogen use | Prescriptions for potential teratogens (including ACE inhibitors and certain anti-emetics) in the 12 months before pregnancy using the Therapy file |
| Eating disorders | Evidence of eating disorders (indication for antidepressants) ever before the start of pregnancy using Read codes in the Clinical and Referral files and ICD-10 codes in HES Diagnosis data, where available |
| Pain disorders | Evidence of pain disorders (indication for antidepressants) ever before the start of pregnancy using Read codes in the Clinical and Referral files and ICD-10 codes in HES Diagnosis data, where available |
| Migraine prophylaxis | Evidence of migraine prophylaxis (indication for antidepressants) ever before the start of pregnancy using Read codes in the Clinical and Referral files and ICD-10 codes in HES Diagnosis data, where available |
| Tension-type headache | Evidence of tension-type headache (indication for antidepressants) ever before the start of pregnancy using Read codes in the Clinical and Referral files and ICD-10 codes in HES Diagnosis data, where available |
| Stress incontinence | Evidence of stress incontinence (indication for antidepressants) ever before the start of pregnancy using Read codes in the Clinical and Referral files and ICD-10 codes in HES Diagnosis data, where available |

^1^ Codelists available via https://github.com/flozoemartin/codelists

#### 2.6 Analysis

##### 2.6.1 Primary analysis

In order to get achieve better exchangeability between exposed and unexposed, we wanted to utilize the depression scale data in CPRD that would help us identify the severity of illness among those who were depressed and/or anxious. There are several depression and anxiety scales administered in primary care in the UK and each follow their own scoring and threshold system to denote mild, moderate, and severe depression. Scale administration was abstracted from the Clinical and Referral data in CPRD GOLD and scores were obtained from the Additional Clinical Details. Scores were then standardized into “none”, “mild”, “moderate”, and “severe” as per the respective scale documentation, summarized in Table S3.

Table S3 Scales and their respective score standardisation breakdown.

| **Scale** | **Score** | | | |
| --- | --- | --- | --- | --- |
|  | **None** | **Mild** | **Moderate** | **Severe** |
| Beck Depression Inventory second edition (BDI-II)^23^ | 0 – 13 | 14 – 19 | 20 – 28 | ≥29 |
| Clinical Outcomes in Routine Evaluation (CORE-10)^24^ | 0 – 5 | 6 – 10 | 15 – 19 | ≥20 |
| Edinburgh Postnatal Depression Scale (EPDS)^25^ | 0 – 6 | 7 – 13 | 14 – 19 | ≥20 |
| Generalised Anxiety Disorder Assessment (GAD-7)^26^ | 0 – 4 | 5 – 9 | 10 – 14 | ≥15 |
| Hospital Anxiety and Depression Scale (HADS)^27^ | 0 – 4 | 5 – 9 | 10 – 14 | ≥15 |
| Hamilton Rating Scale for Depression (Ham-D)^28^ | 0 – 7 | 8 – 16 | 17 – 23 | ≥24 |
| Montgomery-Åsberg Depression Rating Scale (MADRS)^29^ | 0 – 6 | 7 – 19 | 20 – 34 | ≥35 |
| Patient Health Questionnaire (PHQ-9)^27^ | 0 – 4 | 5 – 9 | 10 – 14 | ≥15 |

We then identified the patients who had been administered a scale in the 12 months prior to pregnancy; in order to stratify the primary analysis by those who had been severely depressed in the year before pregnancy, we categorized those who had been administered a scale in the 12 months prior to pregnancy as the highest score (none, mild, moderate, or severe) that they had recorded in that time. We then re-ran the primary analysis, comparing the risk of miscarriage between exposed and unexposed to antidepressants in trimester one, among those who had been identified as severely depressed in the 12 months before pregnancy. Not only are these patients more likely to be exchangeable because they had been administered a scale (which is likely driven by several confounding factors) but also had been deemed to have actively severe depression pre-pregnancy.

##### 2.6.2 Exposure discordant pregnancy analysis

For the exposure discordant analysis, patients with only one pregnancy or multiple but concordant pregnancies (where all their pregnancies were either exposed or unexposed) in the Pregnancy Register were dropped from the analytical sample. Then, discordant pregnancies were compared using multivariable Cox proportional hazards models, generating stratified estimates by patient identifier.

##### 2.6.3 Propensity score matched analysis

As detailed in Desai *et al*., we used their stepwise approach to construct and perform the propensity score analysis.^30^ In the first step, we specified our propensity score model using logistic regression including the covariates shown in Figure 1 on antidepressant use during pregnancy. Having predicted the odds using the output from the regression model, we took the inverse probability as the propensity score for each patient.

Overlap in the propensity score distribution between exposed and unexposed was then assessed. As shown in Figure S2, S4, and S6, there was sufficient overlap to trim non-overlapping regions.

We deemed the target estimand in this study to be the average treatment effect among the treated (ATT), given that this is a drug safety study among a vulnerable population, therefore not all patients who meet the study inclusion criteria would be considered for treatment.^30^ Although not highlighted by Desai *et al.*, similarly to weighting, propensity score matching also facilitates the estimation of the ATT because only those who are treated are matched with comparators.^31^ Given that propensity score matching performed well in comparison to the weighting approaches in the comparison performed by Desai *et al.*, our ability to estimate the ATT, and the use of multiple approaches in triangulation in this study, we chose to move forward with propensity score matching.

Using the R package ‘matchit’, we first matched 10-to-one, nearest neighbour (having discard patients outside of the overlap as this region was deemed to be sufficient) which achieved improved balance between adjusted and unadjusted (Figure S2) (N=129 646 matched, N=91 991 unmatched controls, and N=2 discarded treated). Given that the variables whose standardized mean difference were furthest from zero (Figure S3) were likely associated with healthcare contact (other prescriptions, mental health diagnoses, etc.), we tried exact matching on CPRD consultation event number in the year before pregnancy.

Figure S2 Balance plot for the first matching iteration.

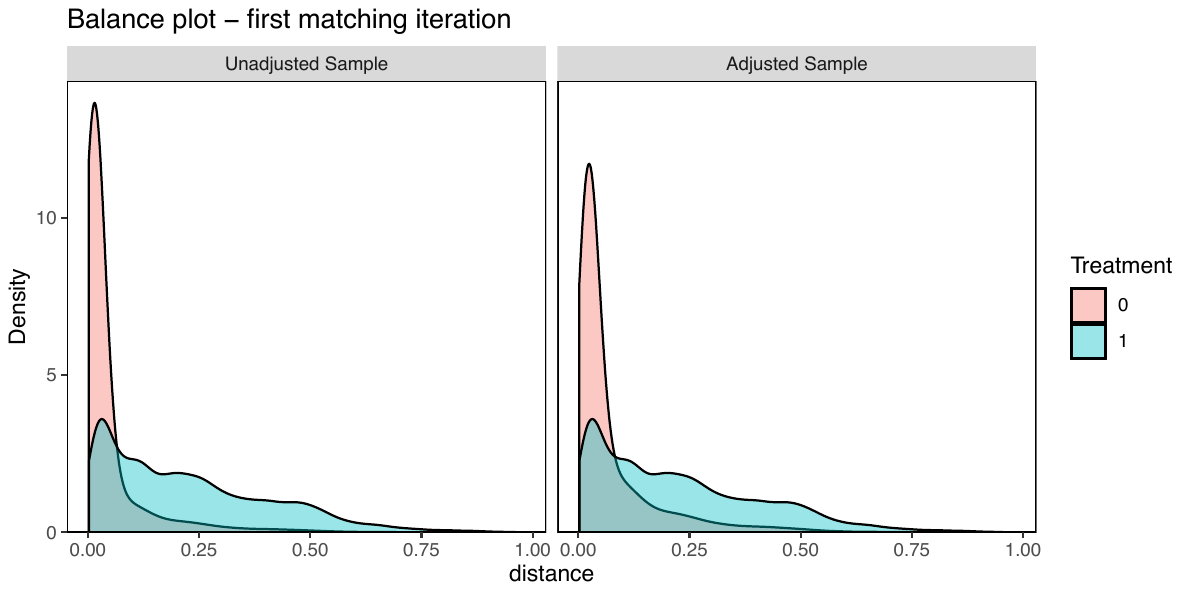

Figure S3 Love plot for the first matching iteration.

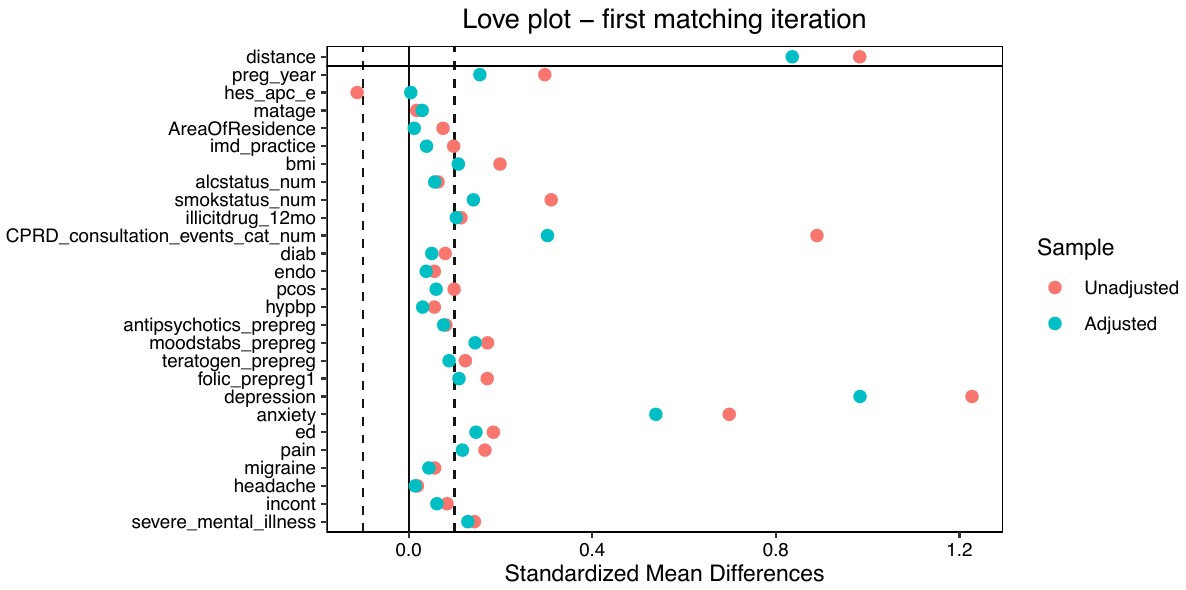

Exact matching on CPRD consultation event number in the year prior to pregnancy helped achieve good balance, having sacrificed number of control matches down to 5 due to insufficient numbers on which to exact match (Figure S4) and smaller standardized mean differences for some more common covariates (like mental health diagnoses), but less so for rarer covariates (such as illicit drug use or antipsychotic use in the 12 months prior to pregnancy) (Figure S5) (N=70 716 matched, N=150 921 unmatched controls, and N=2 discarded treated).

Figure S4 Balance plot for the second matching iteration.

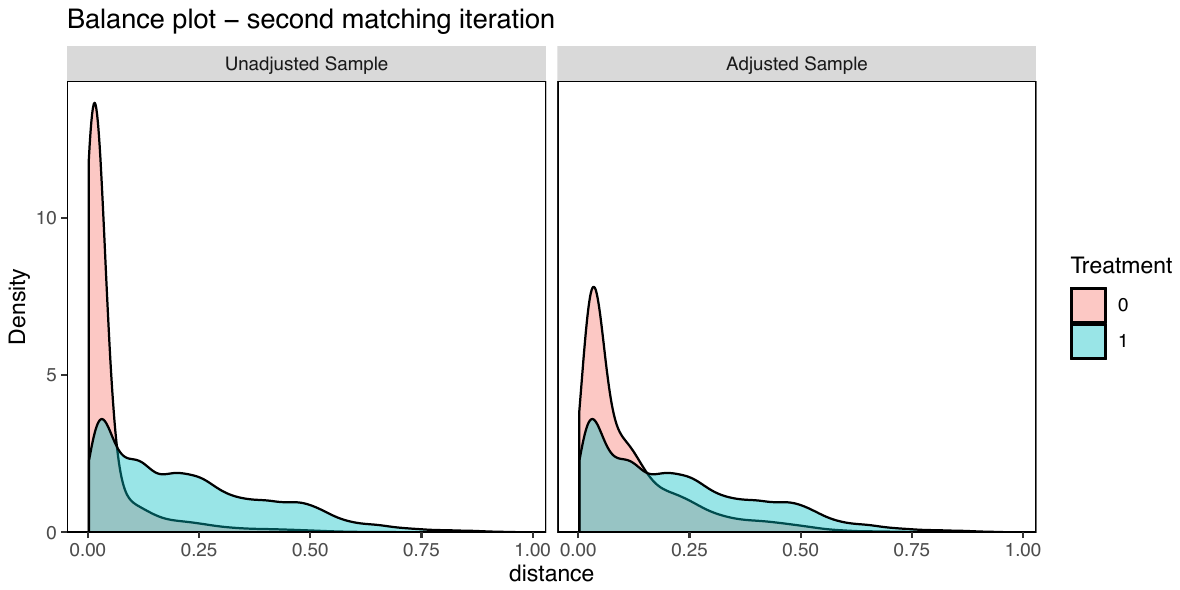

Figure S5 Love plot for the second matching iteration.

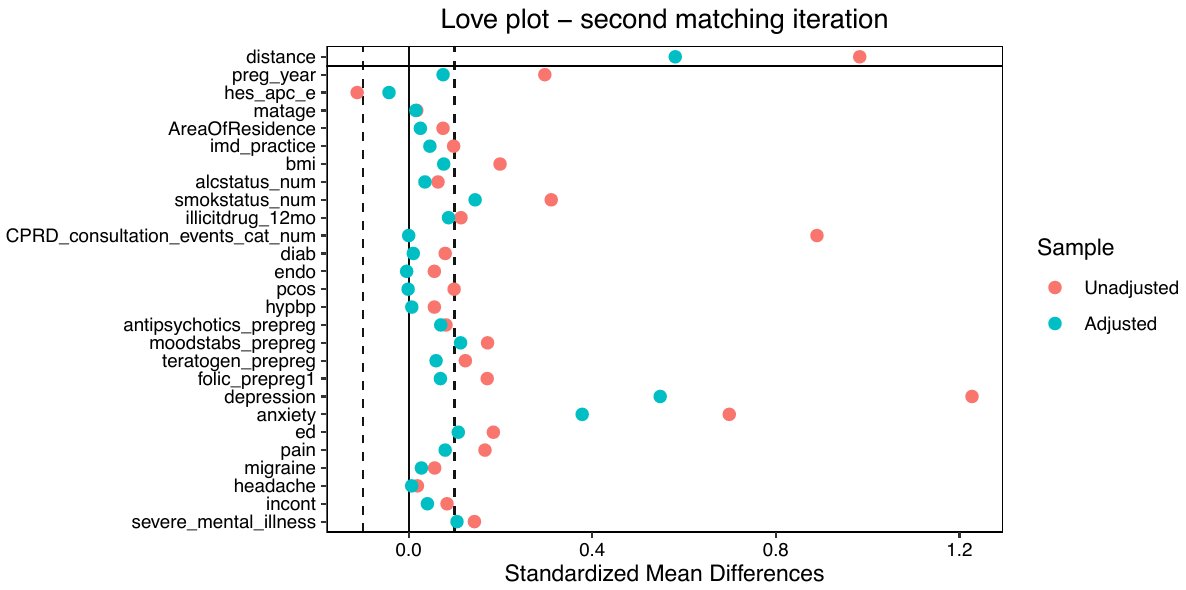

In the third matching iteration, we added a caliper of 0.2, meaning that in order to be matched, patients’ propensity scores had to be within 0.2 SD of the distance measure. This resulted in a slightly higher number of discards but achieved better balance in the adjusted sample (Figure S6 and Figure S7) but precluded matching more than one control to each case (N=23 346 matched, N=198 178 unmatched controls, N=113 unmatched treated, and N=2 discarded treated). We decided to use these matching criteria for the propensity score matched analysis.

Figure S6 Balance plot for the third matching iteration.

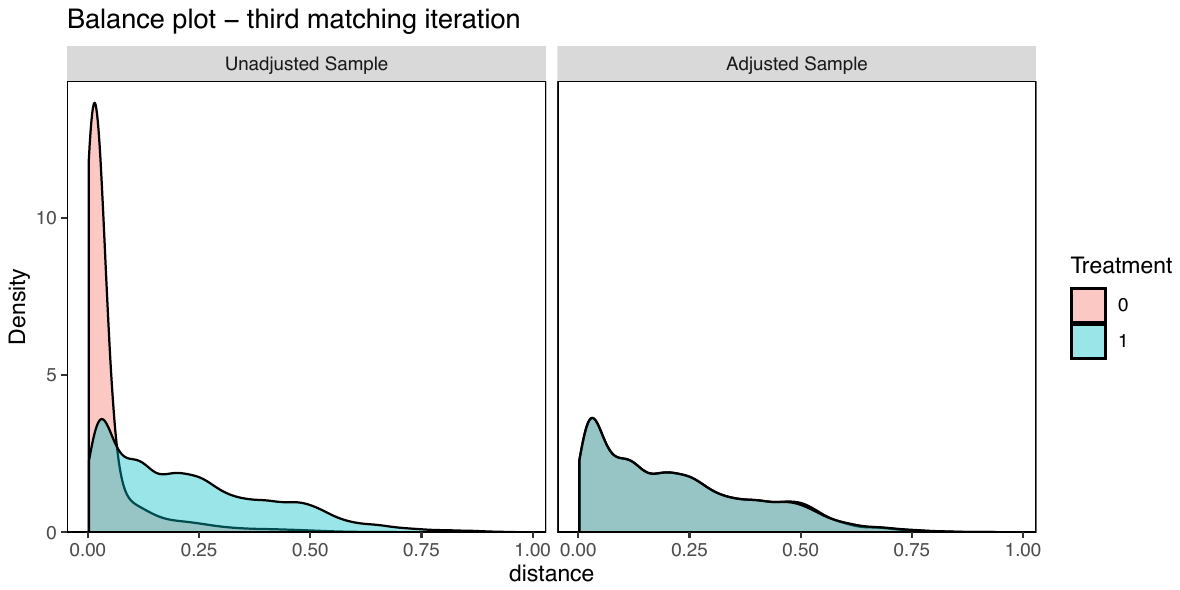

Figure S7 Love plot for the third matching iteration.

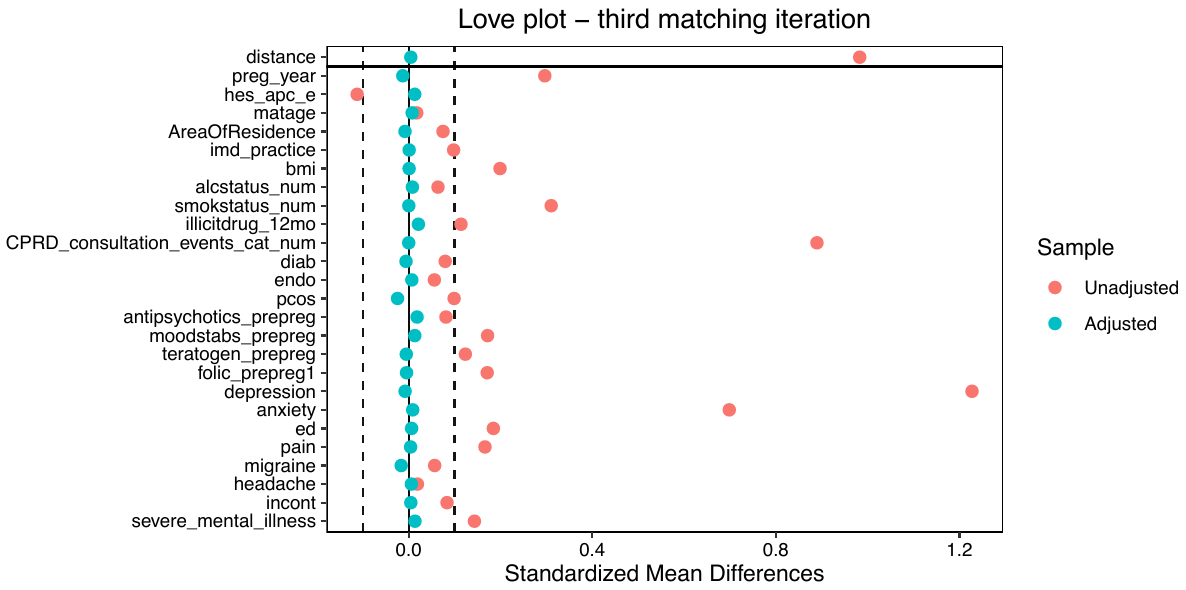

### 3 Results

Table S4 The number of each outcome in the eligible sample, stratified by exposure to antidepressants.

| **Outcome** | **Total (%)** | **Exposed (%)** | **Unexposed (%)** |
| --- | --- | --- | --- |
| Total | 1,021,384 (100) | 73,540 (100) | 947,844 (100) |
| Live birth | 722,672 (70.75) | 45,844 (62.34) | 676,828 (71.41) |
| Stillbirth | 3,140 (0.31) | 241 (0.33) | 2,899 (0.31) |
| Live birth or stillbirth | 511 (0.05) | 44 (0.06) | 467 (0.05) |
| Miscarriage | 127,517 (12.48) | 10,761 (14.63) | 116,756 (12.32) |
| Termination of pregnancy (TOP) | 39,742 (3.89) | 3,986 (5.42) | 35,756 (3.77) |
| Probable TOP | 99,841 (9.78) | 10,427 (14.18) | 89,414 (9.43) |
| Ectopic | 11,157 (1.09) | 952 (1.29) | 10,205 (1.08) |
| Molar | 975 (0.10) | 75 (0.10) | 900 (0.09) |
| Blighted ovum | 738 (0.07) | 62 (0.08) | 676 (0.07) |
| Unspecified loss | 6,495 (0.64) | 554 (0.75) | 5,941 (0.63) |
| Delivery based on a third trimester pregnancy pregnancy record | 6,626 (0.65) | 470 (0.64) | 6,156 (0.65) |
| Delivery based on a late pregnancy record | 1,970 (0.19) | 124 (0.17) | 1,846 (0.19) |

Table S5 Characteristics of those who were excluded based on having an unknown outcome, compared to those who were eligible for inclusion.

| **Variable** | **All** | **Unknown outcome (excluded)** | **Any other outcome (included)** |
| --- | --- | --- | --- |
|  | 1,232,496 (100.0) | 211,112 (100.0) | 1,021,384 (100.0) |
| **Maternal age at start of pregnancy** |  |  |  |
| <18 | 46,200 (3.7) | 7,510 (3.6) | 38,690 (3.8) |
| 18-24 | 280,739 (22.8) | 47,825 (22.7) | 232,914 (22.8) |
| 25-29 | 315,184 (25.6) | 52,142 (24.7) | 263,042 (25.8) |
| 30-34 | 338,652 (27.5) | 55,579 (26.3) | 283,073 (27.7) |
| >=35 | 251,721 (20.4) | 48,056 (22.8) | 203,665 (19.9) |
| **Year of pregnancy** |  |  |  |
| 1996-2000 | 141,308 (11.5) | 22,296 (10.6) | 119,012 (11.7) |
| 2001-2005 | 285,694 (23.2) | 43,408 (20.6) | 242,286 (23.7) |
| 2006-2010 | 369,707 (30.0) | 60,315 (28.6) | 309,392 (30.3) |
| 2011-2015 | 316,169 (25.7) | 59,923 (28.4) | 256,246 (25.1) |
| 2016-2018 | 119,618 (9.7) | 25,170 (11.9) | 94,448 (9.2) |
| **Practice IMD (in quintiles)** |  |  |  |
| 1 | 198,431 (16.1) | 36,938 (17.5) | 161,493 (15.8) |
| 2 | 197,713 (16.0) | 32,122 (15.2) | 165,591 (16.2) |
| 3 | 226,134 (18.3) | 38,964 (18.5) | 187,170 (18.3) |
| 4 | 278,396 (22.6) | 49,187 (23.3) | 229,209 (22.4) |
| 5 | 331,822 (26.9) | 53,901 (25.5) | 277,921 (27.2) |
| **Maternal ethnicity** |  |  |  |
| White | 739,370 (60.0) | 107,756 (51.0) | 631,614 (61.8) |
| South Asian | 38,861 (3.2) | 7,367 (3.5) | 31,494 (3.1) |
| Black | 21,503 (1.7) | 4,797 (2.3) | 16,706 (1.6) |
| Other | 13,938 (1.1) | 2,811 (1.3) | 11,127 (1.1) |
| Mixed | 8,150 (0.7) | 1,561 (0.7) | 6,589 (0.6) |
| Missing | 410,674 (33.3) | 86,820 (41.1) | 323,854 (31.7) |
| **Maternal body mass index (BMI)** |  |  |  |
| Underweight (<18.5 kg/m^2) | 40,432 (3.3) | 6,816 (3.2) | 33,616 (3.3) |
| Healthy weight (18.5-24.9 kg/m^2) | 557,430 (45.2) | 92,320 (43.7) | 465,110 (45.5) |
| Overweight (25.0-29.9 kg/m^2) | 286,244 (23.2) | 47,995 (22.7) | 238,249 (23.3) |
| Obese (>=30.0 kg/m^2) | 216,506 (17.6) | 36,806 (17.4) | 179,700 (17.6) |
| Missing | 131,884 (10.7) | 27,175 (12.9) | 104,709 (10.3) |
| **Maternal history of miscarriage** |  |  |  |
| Yes | 189,375 (15.4) | 28,381 (13.4) | 160,994 (15.8) |
| **Maternal history of stillbirth** |  |  |  |
| Yes | 7,451 (0.6) | 1,147 (0.5) | 6,304 (0.6) |
| **Maternal parity at the start of pregnancy** |  |  |  |
| 0 | 588,472 (47.7) | 102,697 (48.6) | 485,775 (47.6) |
| 1 | 410,740 (33.3) | 64,492 (30.5) | 346,248 (33.9) |
| 2 | 162,235 (13.2) | 30,879 (14.6) | 131,356 (12.9) |
| >=3 | 71,049 (5.8) | 13,044 (6.2) | 58,005 (5.7) |
| **Maternal indications for antidepressants ever before pregnancy** |  |  |  |
| Depression | 305,151 (24.8) | 52,795 (25.0) | 252,356 (24.7) |
| Anxiety | 186,333 (15.1) | 31,939 (15.1) | 154,394 (15.1) |
| **Maternal severe mental illness ever before the start of pregnancy** |  |  |  |
| Yes | 6,371 (0.5) | 1,292 (0.6) | 5,079 (0.5) |
| **Number of consultations in the 12 months before pregnancy** |  |  |  |
| 0 | 144,082 (11.7) | 25,030 (11.9) | 119,052 (11.7) |
| 1-3 | 305,799 (24.8) | 43,658 (20.7) | 262,141 (25.7) |
| 4-10 | 501,172 (40.7) | 81,421 (38.6) | 419,751 (41.1) |
| >10 | 281,443 (22.8) | 61,003 (28.9) | 220,440 (21.6) |
| **Smoking status around the start of pregnancy** |  |  |  |
| Non-smoker | 502,551 (40.8) | 87,788 (41.6) | 414,763 (40.6) |
| Current smoker | 368,611 (29.9) | 63,714 (30.2) | 304,897 (29.9) |
| Ex-smoker | 295,170 (23.9) | 46,905 (22.2) | 248,265 (24.3) |
| Missing | 66,164 (5.4) | 12,705 (6.0) | 53,459 (5.2) |
| **Other prescriptions 12 months before pregnancy** |  |  |  |
| Antipsychotics | 1,076 (0.1) | 211 (0.1) | 865 (0.1) |
| Mood stabilisers | 12,576 (1.0) | 2,664 (1.3) | 9,912 (1.0) |
| Folic acid (5mg) | 67,479 (5.5) | 8,649 (4.1) | 58,830 (5.8) |

Table S6 Findings from the primary and secondary analysis (Figure 2) with total contributed days of follow-up.

|  | **Total** | **n/N (%)** | **Total contributed time (days)** | **HR (95%CI)** | **aHR (95%CI)** |
| --- | --- | --- | --- | --- | --- |
| **Primary analyses** | | | | | |
| **Primary Cox model** | | | | | |
| Unexposed in T1 | 979,169 | 111,315/907,756 (12.3) | 129,353,056 | 1.00 (ref) | 1.00 (ref) |
| Exposed in T1 |  | 10,442/71,413 (14.6) | 9,672,654 | 1.21 (1.19-1.23) | 1.04 (1.02-1.06) |
| **Exposure discordant analysis** | | | | | |
| Unexposed in T1 | 78,072 | 6,264/ 47,737 (13.1) | 5,927,538 | 1.00 (ref) | 1.00 (ref) |
| Exposed in T1 |  | 5,034/30,335 (16.6) | 4,052,565 | 1.19 (1.15-1.24) | 1.20 (1.16-1.25) |
| **Propensity score matched analysis** | | | | | |
| Unexposed in T1 | 25,026 | 1,556/ 13,354 (11.7) | 1,732,968 | 1.00 (ref) | 1.00 (ref) |
| Exposed in T1 |  | 1,712/11,672 (14.7) | 1,621,430 | - | 1.09 (1.02-1.17) |
| **Secondary analyses** | | | | | |
| **Pattern analysis** | | | | | |
| Unexposed in T1 | 979,169 | 111,315/907,756 (12.3) | 129,353,056 | 1.00 (ref) | 1.00 (ref) |
| Prevalent, exposed in T1 |  | 8,751/60,167 (14.5) | 8,198,643 | 1.19 (1.17-1.22) | 1.00 (0.98-1.03) |
| Incident, exposed in T1 |  | 1,691/11,246 (15.0) | 1,474,011 | 1.29 (1.23-1.35) | 1.24 (1.19-1.30) |
| **Class analysis** | | | | | |
| Unexposed in T1 | 979,169 | 111,315/907,756 (12.3) | 129,353,056 | 1.00 (ref) | 1.00 (ref) |
| SSRI exposed in T1 |  | 7,428/52,244 (14.2) | 7,072,641 | 1.18 (1.15-1.20) | 1.03 (1.00-1.05) |
| SNRI exposed in T1 |  | 587/3,658 (16.0) | 484,332 | 1.33 (1.23-1.44) | 1.04 (0.96-1.12) |
| TCA exposed in T1 |  | 1,255/8,205 (15.3) | 1,134,021 | 1.26 (1.20-1.33) | 1.06 (1.01-1.12) |
| Other exposed in T1 |  | 509/3,104 (16.4) | 411,173 | 1.36 (1.25-1.48) | 1.13 (1.04-1.23) |
| Multiple in T1 |  | 663/4,202 (15.8) | 570,487 | 1.30 (1.21-1.40) | 1.03 (0.96-1.11) |
| **Dose analysis** | | | | | |
| Unexposed in T1 | 979,169 | 111,315/907,756 (12.3) | 129353056 | 1.00 (ref) | 1.00 (ref) |
| Low dose in T1 |  | 4,394/31,214 (14.1) | 4,246,387 | 1.17 (1.13-1.20) | 1.05 (1.01-1.08) |
| Medium dose in T1 |  | 4,738/31,416 (15.1) | 4,239,483 | 1.25 (1.21-1.28) | 1.05 (1.02-1.08) |
| High dose in T1 |  | 1,310/8,783 (14.9) | 1,186,784 | 1.23 (1.16-1.30) | 0.97 (0.91-1.02) |

Table S7 More stringent definitions of the exposure: for the primary analysis, requiring at least two prescriptions during pregnancy to be considered exposed.

|  | **Total^1^** | **Miscarriage n/total N (%)** | **Total time (days)** | **HR** | **aHR^2^** |
| --- | --- | --- | --- | --- | --- |
| **Primary Cox** | | | | | |
| Unexposed in T1 | 979,169 | 114,915/933,859 (12.3) | 132,932,592 | 1.00 (ref) | 1.00 (ref) |
| Exposed in T1 |  | 6,842/45,310 (15.1) | 6,093,118 | 1.24 (1.21-1.27) | 1.02 (1.00-1.05) |
| **Pattern analysis** | | | | | |
| Unexposed in T1 | 979,169 | 114,915/933,859 (12.3) | 132,932,592 | 1.00 (ref) | 1.00 (ref) |
| Prevalent use in T1 |  | 6,279/41,780 (15.0) | 5,641,582 | 1.23 (1.20-1.26) | 1.00 (0.98-1.03) |
| Incident use in T1 |  | 563/3,530 (15.9) | 451,536 | 1.32 (1.22-1.42) | 1.27 (1.17-1.37) |

^1^ Total includes episodes that were exposed, unexposed and exposed who contributed unexposed time, in other words trimester one initiators

^2^ Adjusted for maternal age at the start of pregnancy, IMD quintile, year of pregnancy, history of miscarriage, smoking around the start of pregnancy, parity at the start of pregnancy, use of high dose folic acid, antipsychotics, and mood stabilisers in the 12 months before pregnancy, number of primary care consultations in the 12 months before pregnancy and depression or anxiety ever before the start of pregnancy

Table S8 Indication-based sample sensitivity analyses: primary and pattern analysis among those with depression or anxiety noted in the 12 months prior to pregnancy and the primary analysis among those with ‘severe’ depression or anxiety as measured by the scales summarized in Table S2.

|  | **Total^1^** | **Miscarriage n/total N (%)** | **Total exposed time (days)** | **HR (95%CI)** | **aHR^2^ (95%CI)** |
| --- | --- | --- | --- | --- | --- |
| **Primary analysis among those with depression and/or anxiety noted in the 12 months prior to pregnancy** |  |  |  |  |  |
| Unexposed during T1 | 99,820 | 7,646/62,563 (12.22) | 8,475,726 | 1.00 (ref) | 1.00 (ref) |
| Exposed during T1 |  | 5,291/37,257 (14.20) | 5,064,507 | 1.13 (1.09-1.17) | 1.04 (1.01-1.08) |
| **Pattern analysis among those with depression and/or anxiety noted in the 12 months prior to pregnancy** |  |  |  |  |  |
| Unexposed in T1 among actively indicated | 99,820 | 7,646/62,563 (12.2) | 8,475,726 | 1.00 (ref) | 1.00 (ref) |
| Prevalent exposed in T1 among actively indicated |  | 4,898/34,671 (14.1) | 4,726,880 | 1.12 (1.08-1.16) | 1.03 (0.99-1.07) |
| Incident exposed in T1 among actively indicated |  | 393/2,586 (15.2) | 337,627 | 1.25 (1.13-1.37) | 1.24 (1.13-1.37) |
| **Primary analysis among those with “severe depression”** **and/or anxiety in the 12 months prior to pregnancy^3^** |  |  |  |  |  |
| Unexposed during T1 | 9,170 | 656/5,044 (13.01) | 674,575 | 1.00 (ref) | 1.00 (ref) |
| Exposed during T1 |  | 588/4,126 (14.25) | 558,676 | 1.06 (0.95-1.18) | 1.02 (0.92-1.14) |

^1^ Total includes episodes that were exposed, unexposed and exposed who contributed unexposed time, in other words trimester one initiators

^2^ Adjusted for maternal age at the start of pregnancy, IMD quintile, year of pregnancy, history of miscarriage, smoking around the start of pregnancy, parity at the start of pregnancy, use of high dose folic acid, antipsychotics, and mood stabilisers in the 12 months before pregnancy, and number of primary care consultations in the 12 months before pregnancy

^3^ Severity scoring described in Table S3

Table S9 Negative control-type analysis comparing ‘prevalent’ users to those who discontinued in the 3 months prior to pregnancy.

|  | **Total** | **Miscarriage n/total N (%)** | **Total contributed time (days)** | **HR (95%CI)** | **aHR^1^ (95%CI)** |
| --- | --- | --- | --- | --- | --- |
| **Negative control-type analysis** | | | | | |
| Unexposed in T1 (discontinued) | 84,577 | 3,206/24,410 (13.13) | 3,412,069 | 1.00 (ref) | 1.00 (ref) |
| Exposed in T1 |  | 8,751/60,167 (14.54) | 8,198,643 | 1.12 (1.07-1.16) | 1.00 (0.97-1.04) |

^1^ Adjusted for maternal age at the start of pregnancy, IMD quintile, year of pregnancy, history of miscarriage, smoking around the start of pregnancy, parity at the start of pregnancy, use of high dose folic acid, antipsychotics, and mood stabilisers in the 12 months before pregnancy, and number of primary care consultations in the 12 months before pregnancy

Table S10 Exposure discordant pregnancy sensitivity analysis, stratifying by exposure discordant groups where the first pregnancy is exposed and subsequent pregnancies are not, then where the first pregnancy is not exposed and subsequent pregnancies in the group are.

|  | **Total** | **n/N (%)** | **Total contributed time (days)** | **HR (95%CI)** | **aHR (95%CI)** |
| --- | --- | --- | --- | --- | --- |
| **First pregnancy in group exposed** | | | | | |
| Unexposed in T1 | 18,111 | 1,273/ 10,120 (12.6) | 1,285,886 | 1.00 (ref) | 1.00 (ref) |
| Exposed in T1 (first pregnancy) |  | 1,698/ 7,991 (21.2) | 1,052,708 | 1.68 (1.56-1.80) | 1.98 (1.82-2.16) |
| **Subsequent pregnancy in group exposed** | | | | | |
| Unexposed in T1 | 59,961 | 4,991/ 37,617 (13.3) | 4,641,652 | 1.00 (ref) | 1.00 (ref) |
| Exposed in T1 (subsequent pregnancy) |  | 3,336/22,344 (14.9) | 2,999,857 | 1.05 (1.01-1.10) | 0.97 (0.93-1.02) |

Table S11 Characteristics of prevalent and incident users of antidepressants during trimester one, as well as unexposed.

| Variable | **All** | **Incident, exposed in T1** | **Prevalent, exposed in T1** | **Unexposed in T1** |
| --- | --- | --- | --- | --- |
|  | 1,021,384 (100.0) | 11,822 (100.0) | 61,718 (100.0) | 947,844 (100.0) |
| **Maternal age at start of pregnancy** |  |  |  |  |
| <18 | 38,690 (3.8) | 332 (2.8) | 667 (1.1) | 37,691 (4.0) |
| 18-24 | 232,914 (22.8) | 3,989 (33.7) | 13,624 (22.1) | 215,301 (22.7) |
| 25-29 | 263,042 (25.8) | 3,015 (25.5) | 16,064 (26.0) | 243,963 (25.7) |
| 30-34 | 283,073 (27.7) | 2,460 (20.8) | 16,686 (27.0) | 263,927 (27.8) |
| >=35 | 203,665 (19.9) | 2,026 (17.1) | 14,677 (23.8) | 186,962 (19.7) |
| **Year of pregnancy** |  |  |  |  |
| 1996-2000 | 119,012 (11.7) | 1,129 (9.5) | 3,691 (6.0) | 114,192 (12.0) |
| 2001-2005 | 242,286 (23.7) | 2,735 (23.1) | 11,967 (19.4) | 227,584 (24.0) |
| 2006-2010 | 309,392 (30.3) | 3,246 (27.5) | 17,095 (27.7) | 289,051 (30.5) |
| 2011-2015 | 256,246 (25.1) | 3,231 (27.3) | 19,348 (31.3) | 233,667 (24.7) |
| 2016-2018 | 94,448 (9.2) | 1,481 (12.5) | 9,617 (15.6) | 83,350 (8.8) |
| **Practice IMD (in quintiles)** |  |  |  |  |
| 1 (least deprived) | 161,493 (15.8) | 1,403 (11.9) | 8,157 (13.2) | 151,933 (16.0) |
| 2 | 165,591 (16.2) | 1,712 (14.5) | 9,313 (15.1) | 154,566 (16.3) |
| 3 | 187,170 (18.3) | 2,053 (17.4) | 11,279 (18.3) | 173,838 (18.3) |
| 4 | 229,209 (22.4) | 2,833 (24.0) | 14,247 (23.1) | 212,129 (22.4) |
| 5 (most deprived) | 277,921 (27.2) | 3,821 (32.3) | 18,722 (30.3) | 255,378 (26.9) |
| **Maternal ethnicity** |  |  |  |  |
| White | 631,614 (61.8) | 7,631 (64.5) | 40,035 (64.9) | 583,948 (61.6) |
| South Asian | 31,494 (3.1) | 200 (1.7) | 665 (1.1) | 30,629 (3.2) |
| Black | 16,706 (1.6) | 130 (1.1) | 340 (0.6) | 16,236 (1.7) |
| Other | 11,127 (1.1) | 80 (0.7) | 244 (0.4) | 10,803 (1.1) |
| Mixed | 6,589 (0.6) | 82 (0.7) | 307 (0.5) | 6,200 (0.7) |
| Missing | 323,854 (31.7) | 3,699 (31.3) | 20,127 (32.6) | 300,028 (31.7) |
| **Maternal body mass index (BMI)** |  |  |  |  |
| Underweight (<18.5 kg/m^2^) | 33,616 (3.3) | 514 (4.3) | 2,186 (3.5) | 30,916 (3.3) |
| Healthy weight (18.5-24.9 kg/m^2^) | 465,110 (45.5) | 4,835 (40.9) | 24,012 (38.9) | 436,263 (46.0) |
| Overweight (25.0-29.9 kg/m^2^) | 238,249 (23.3) | 2,628 (22.2) | 14,775 (23.9) | 220,846 (23.3) |
| Obese (>=30.0 kg/m^2^) | 179,700 (17.6) | 2,600 (22.0) | 15,940 (25.8) | 161,160 (17.0) |
| Missing | 104,709 (10.3) | 1,245 (10.5) | 4,805 (7.8) | 98,659 (10.4) |
| **Maternal history of miscarriage** |  |  |  |  |
| Yes | 160,994 (15.8) | 2,284 (19.3) | 12,129 (19.7) | 146,581 (15.5) |
| **Maternal history of stillbirth** |  |  |  |  |
| Yes | 6,304 (0.6) | 94 (0.8) | 592 (1.0) | 5,618 (0.6) |
| **Maternal parity at the start of pregnancy** |  |  |  |  |
| 0 | 485,775 (47.6) | 4,480 (37.9) | 22,746 (36.9) | 458,549 (48.4) |
| 1 | 346,248 (33.9) | 3,969 (33.6) | 20,729 (33.6) | 321,550 (33.9) |
| 2 | 131,356 (12.9) | 2,145 (18.1) | 11,673 (18.9) | 117,538 (12.4) |
| >=3 | 58,005 (5.7) | 1,228 (10.4) | 6,570 (10.6) | 50,207 (5.3) |
| **Maternal indications for antidepressants ever before the end of pregnancy** |  |  |  |  |
| Depression | 252,356 (24.7) | 6,376 (53.9) | 49,929 (80.9) | 196,051 (20.7) |
| Anxiety | 154,394 (15.1) | 3,777 (31.9) | 30,064 (48.7) | 120,553 (12.7) |
| **Maternal severe mental illness ever before the start of pregnancy** |  |  |  |  |
| Yes | 5,079 (0.5) | 165 (1.4) | 1,609 (2.6) | 3,305 (0.3) |
| **Number of consultations in the 12 months before the end of pregnancy** |  |  |  |  |
| 0 | 119,052 (11.7) | 984 (8.3) | 3,992 (6.5) | 114,076 (12.0) |
| 1-3 | 262,141 (25.7) | 1,617 (13.7) | 3,048 (4.9) | 257,476 (27.2) |
| 4-10 | 419,751 (41.1) | 4,570 (38.7) | 19,841 (32.1) | 395,340 (41.7) |
| >10 | 220,440 (21.6) | 4,651 (39.3) | 34,837 (56.4) | 180,952 (19.1) |
| **Smoking status around the start of pregnancy** |  |  |  |  |
| Non-smoker | 414,763 (40.6) | 3,176 (26.9) | 16,749 (27.1) | 394,838 (41.7) |
| Current smoker | 304,897 (29.9) | 5,532 (46.8) | 26,596 (43.1) | 272,769 (28.8) |
| Ex-smoker | 248,265 (24.3) | 2,585 (21.9) | 16,822 (27.3) | 228,858 (24.1) |
| Missing | 53,459 (5.2) | 529 (4.5) | 1,551 (2.5) | 51,379 (5.4) |
| **Other prescriptions 12 months before pregnancy** |  |  |  |  |
| Antipsychotics | 865 (0.1) | 63 (0.5) | 434 (0.7) | 368 (0.0) |
| Mood stabilisers | 9,912 (1.0) | 275 (2.3) | 2,726 (4.4) | 6,911 (0.7) |
| Folic acid (5mg) | 58,830 (5.8) | 864 (7.3) | 6,308 (10.2) | 51,658 (5.5) |

Table S12 Main and secondary analyses restricted to those with linked data.

|  | **Total^1^** | **Miscarriage n/total N (%)** | **Total contributed time (in days)** | **HR (95% CI)** | **aHR^2^ (95% CI)** |
| --- | --- | --- | --- | --- | --- |
| **Primary analyses** | | | | | |
| **Primary Cox model** | | | | | |
| Unexposed in T1 | 473,736 | 54,150/443,545 (12.2) | 63,431,124 | 1.00 (ref) | 1.00 (ref) |
| Exposed in T1 |  | 4,359/30,191 (14.4) | 4,094,862 | 1.20 (1.17-1.24) | 1.03 (1.00-1.06) |
| **Exposure discordant analysis** | | | | | |
| Unexposed in T1 | 34,465 | 2,748/21,271 (12.9) | 2,626,691 | 1.00 (ref) | 1.00 (ref) |
| Exposed in T1 |  | 2,131/13,194 (16.2) | 1,761,377 | 1.18 (1.11-1.24) | 1.18 (1.12-1.26) |
| **Propensity score matched^3^ analysis** | | | | | |
| Unexposed in T1 | 24,242 | 2,392/19,184 (12.5) | 2,682,273 | 1.00 (ref) | 1.00 (ref) |
| Exposed in T1 |  | 747/5,058 (14.8) | 700,233 | - | 1.08 (0.99-1.17) |
| **Secondary analyses** | | | | | |
| **Pattern analysis** | | | | | |
| Unexposed in T1 | 473,736 | 54,150/443,545 (12.2) | 63,431,124 | 1.00 (ref) | 1.00 (ref) |
| Prevalent, exposed in T1 |  | 3,617/24,977 (14.5) | 3,409,223 | 1.20 (1.16-1.24) | 1.00 (0.96-1.03) |
| Incident, exposed in T1 |  | 742/5,214 (14.2) | 685,639 | 1.24 (1.15-1.33) | 1.21 (1.13-1.30) |
| **Class analysis** | | | | | |
| Unexposed in T1 | 473,736 | 54,150/443,545 (12.2) | 63,431,124 | 1.00 (ref) | 1.00 (ref) |
| SSRI exposed in T1 |  | 3,097/22,348 (13.9) | 3,026,868 | 1.16 (1.12-1.20) | 1.01 (0.97-1.05) |
| TCA exposed in T1 |  | 610/3,939 (15.5) | 545,470 | 1.29 (1.20-1.40) | 1.08 (1.00-1.17) |
| SNRI exposed in T1 |  | 240/1,340 (17.9) | 174,930 | 1.50 (1.33-1.70) | 1.15 (1.03-1.30) |
| Other exposed in T1 |  | 149/897 (16.6) | 119,405 | 1.39 (1.19-1.62) | 1.12 (0.96-1.30) |
| Multiple in T1^4^ |  | 263/1,667 (15.8) | 228,189 | 1.31 (1.17-1.48) | 1.03 (0.92-1.16) |
| **Dose analysis** | | | | | |
| Unexposed in T1 | 473,736 | 54,150/443,545 (12.2) | 63,431,124 | 1.00 (ref) | 1.00 (ref) |
| Low dose in T1 |  | 1,933/13,994 (13.8) | 1,907,162 | 1.15 (1.10-1.21) | 1.03 (0.99-1.08) |
| Medium dose in T1 |  | 1,910/12,656 (15.1) | 1,705,154 | 1.26 (1.20-1.32) | 1.05 (1.01-1.10) |
| High dose in T1 |  | 516/3,541 (14.6) | 482,546 | 1.21 (1.11-1.32) | 0.94 (0.86-1.02) |

^1^ Total includes episodes that were exposed, unexposed and exposed who contributed unexposed time, in other words trimester one initiators

^2^ Adjusted for maternal age at the start of pregnancy, IMD quintile, year of pregnancy, history of miscarriage, smoking around the start of pregnancy, parity at the start of pregnancy, use of high dose folic acid, antipsychotics, and mood stabilisers in the 12 months before pregnancy, and number of primary care consultations in the 12 months before pregnancy

^3^ The propensity scores additionally contained whether the woman had linked data, body mass index around the start of pregnancy, area of residence, alcohol use, smoking, and illicit drug use around the start of pregnancy, diabetes, endometriosis, and polycystic ovary syndrome diagnosed prior to the start of pregnancy, potential teratogen prescription in the 12 months prior to pregnancy, and eating disorder, pain disorder, migraine prophylaxis, tension-type headache or stress incontinence noted ever before the start of pregnancy

4 Prescribed >1 class of antidepressants during trimester one

Table S13 Risk of having an unknown outcome pregnancy having had depression or anxiety diagnosed or been prescribed antidepressants in the 12 months prior to pregnancy.

|  | **Total** | **Miscarriage n/total N (%)** | **Unexposed n/N (%)** | **OR** |
| --- | --- | --- | --- | --- |
| Antidepressant use in the 12 months prior to pregnancy | 1,166,332 | 28,863/165,942 (17.4) | 169,544/1,000,390 (16.9) | 1.03 (1.02-1.05) |
| Depression in the 12 months prior to pregnancy | 1,166,332 | 15,041/87,191 (17.3) | 183,366/1,079,141 (17.0) | 1.02 (1.00-1.04) |
| Anxiety in the 12 months prior to pregnancy | 1,166,332 | 7,535/42,559 (17.7) | 190,872/1,123,773 (17.0) | 1.05 (1.02-1.08) |

Table S14 Additional adjustment for covariates with substantial missing data.

|  | **Total^1^** | **Miscarriage n/total N (%)** | **Total contributed time (days)** | **HR** | **aHR^2^** |
| --- | --- | --- | --- | --- | --- |
| **Additionally adjusted primary analysis** | | | | | |
| Unexposed in T1 | 618,616 | 71,661/573,571 (12.49) | 82,082,688 | 1.00 (ref) | 1.00 (ref) |
| Exposed in T1 |  | 6,607/45,045 (14.67) | 6,107,730 | 1.19 (1.16-1.22) | 1.03 (1.01-1.06) |

^1^ Total includes episodes that were exposed, unexposed and exposed who contributed unexposed time, in other words trimester one initiators

^2^ Adjusted for maternal age at the start of pregnancy, IMD quintile, year of pregnancy, history of miscarriage, smoking around the start of pregnancy, parity at the start of pregnancy, use of high dose folic acid, antipsychotics, and mood stabilisers in the 12 months before pregnancy, and number of primary care consultations in the 12 months before pregnancy

Table S15 Association between missingness in the excluded variables from the main analysis adjustment set and the outcome, miscarriage.

|  | **Miscarriage n/total N (%)** | **Delivery n/N (%)** | **OR** | **aOR^1^** |
| --- | --- | --- | --- | --- |
| Ethnicity missingness | 39,680/127,334 (31.16) | 230,634/734,919 (31.38) | 0.99 (0.98-1.00) | 1.04 (1.02-1.05) |
| BMI missingness | 11,560/127,334 (9.08) | 69,266/734,919 (9.42) | 0.96 (0.94-0.98) | 1.11 (1.09-1.13) |
| Smoking missingness | 5,577/127,334 (4.38) | 32,654/734,919 (4.44) | 0.99 (0.96-1.01) | 1.31 (1.27-1.36) |

Table S16 Primary analysis restricted to nulliparous women.

|  | **Total^1^** | **Miscarriage n/total N (%)** | **Total exposed time (days)** | **HR** | **aHR^2^** |
| --- | --- | --- | --- | --- | --- |
| **Restricted to nulliparous** | | | | | |
| Unexposed in T1 | 405,807 | 47,928/382,026 (12.55) | 54,413,804 | 1.00 (ref) | 1.00 (ref) |
| Exposed in T1 |  | 3,724/23,781 (15.66) | 3,243,639 | 1.27 (1.23-1.31) | 1.07 (1.03-1.10) |

^1^ Total includes episodes that were exposed, unexposed and exposed who contributed unexposed time, in other words trimester one initiators

^2^ Adjusted for maternal age at the start of pregnancy, IMD quintile, year of pregnancy, history of miscarriage, smoking around the start of pregnancy, parity at the start of pregnancy, use of high dose folic acid, antipsychotics, and mood stabilisers in the 12 months before pregnancy, and number of primary care consultations in the 12 months before pregnancy

Table S17 Miscarriage includes ectopic pregnancy and molar pregnancy.

|  | **Total^1^** | **Miscarriage n/total N (%)** | **Total exposed time (days)** | **HR** | **aHR^2^** |
| --- | --- | --- | --- | --- | --- |
| **Ectopic and molar pregnancy included in miscarriage** | | | | | |
| Unexposed in T1 | 892,941 | 111,988/826,881 (13.54) | 118,197,664 | 1.00 (ref) | 1.00 (ref) |
| Exposed in T1 |  | 10,635/66,060 (16.10) | 8,964,845 | 1.21 (1.18-1.23) | 1.03 (1.01-1.05) |

^1^ Total includes episodes that were exposed, unexposed and exposed who contributed unexposed time, in other words trimester one initiators

^2^ Adjusted for maternal age at the start of pregnancy, IMD quintile, year of pregnancy, history of miscarriage, smoking around the start of pregnancy, parity at the start of pregnancy, use of high dose folic acid, antipsychotics, and mood stabilisers in the 12 months before pregnancy, and number of primary care consultations in the 12 months before pregnancy
